# When time-dependence in disease outcome risk is not captured by impact evaluation modeling studies: a measles vaccination case study

**DOI:** 10.1101/2021.07.12.21260376

**Authors:** Allison Portnoy, Yuli Lily Hsieh, Kaja Abbas, Petra Klepac, Heather Santos, Logan Brenzel, Mark Jit, Matthew Ferrari

## Abstract

**Background:** In modeling studies that evaluate the effects of health programs, the risk of secondary outcomes attributable to infection can vary with underlying disease incidence. Consequently, the impact of interventions on secondary outcomes would not be proportional to incidence reduction. Here we use a case study on measles vaccine program to demonstrate how failure to capture this non-linear relationship can lead to over- or under-estimation.

**Methods:** We used a published model of measles CFR that depends on incidence and vaccine coverage to illustrate the effects of: (1) assuming higher CFR in “no-vaccination” scenarios; (2) time-varying CFRs over the past; and (3) time-varying CFRs in future projections on measles impact estimation. We evaluated how different assumptions on vaccine coverage, measles incidence, and CFR levels in “no-vaccination” scenarios affect estimation of future deaths averted by measles vaccination.

**Results:** Compared to constant CFRs, aligning both “vaccination” and “no-vaccination” scenarios with time variant measles CFR estimates led to larger differences in mortality in historical years and lower in future years.

**Conclusions:** To assess consequences of interventions, impact estimates should consider the effect of “no-intervention” scenario assumptions on model parameters to project estimated impact for alternative scenarios according to intervention strategies and investment decisions.

## Introduction

Model-based estimation has been widely used to evaluate the impact of infectious disease intervention programs outside of empirical observations [1]. There are myriad policy interests in both retrospective program evaluation to estimate the effect of a previously implemented program and prospective program evaluation to project the impact of different intervention options in the future. In both types of program evaluation, the program impact is often quantified by comparing the estimated effects in the scenario with the intervention against those in a scenario without the intervention (e.g., with and without vaccination). Under this framework, detailed methodological considerations defining both the “intervention” and “no-intervention” scenario are a necessary condition to minimize bias in program effect estimates.

The most common approach in developing the “no-intervention” scenario is to “switch off” the program in the model. In regression models, this “switch off” can be incorporated by including an indicator variable for program implementation. In mechanistic simulation models, it can be modeled by setting the uptake of the intervention to zero in scenarios without program implementation while keeping all other parameters consistent with scenario(s) where the program is implemented. For both types of models, we can evaluate the intervention impact as the difference in the outcome of interest, such as the number of deaths under the intervention and no-intervention scenario. Many health impact models assume risks of infectious disease health outcomes conditional on infection, such as case-fatality ratios (CFRs), are independent of disease incidence and health system characteristics. However, there is evidence that the outcome of infection and consequences of infection can depend on health system burden [2-8].

An example of such an intervention is measles vaccination. Due to limited primary data of measles CFR, for many years, the impact of measles vaccine has been estimated by simulation models that assume constant measles CFR over time [9, 10]. However, a recently updated meta-analysis [5] showed a decreasing trend in measles CFR in low- and middle-income countries (LMICs) and found that this is associated with trends in measles vaccination coverage [11], measles incidence [5], and under-five mortality [12]. In most settings, these factors have varied over time and may continue to change in the future, either continuing their past trends or potentially reversing direction following the COVID-19 pandemic [13, 14].

Consequently, in modeling studies that aim to evaluate the impact of a measles vaccination program, changes in the health system is an important aspect to consider as it is related to the risk of disease outcome. As disease management and health system capacity improves over time, we would expect risks of disease outcome to improve over time. Likewise, changes to the population-level risks of disease outcome could be negatively impacted by changes to preventive measures such as vaccination coverage. These observations motivate the design of this study to evaluate the consequences of more realistic assumptions that affect model predictions in the “vaccination” and “no-vaccination” scenarios, accounting for time-dependent elements.

Using measles as a case study, we propose a methodological innovation to model CFR dynamically, which addresses both: (1) dependence of CFR on incidence and other health system characteristics; (2) calculating impact with due consideration for the “no-vaccination” scenario. This paper is organized into two parts. In Part I, we used a log-linear regression model to evaluate three different sets of scenarios where covariates used to estimate measles CFR can be time variant or time invariant. As a baseline, we used the current practice of assuming time and strategy invariant CFRs as described in Wolfson, *et al*. [10]. We also considered two other scenarios in which CFRs depend on covariates that change over time, as informed by our previous study where we showed that CFRs have changed over time from 1980–2006 [5]. First, we only allowed for CFRs to change over past time according to the log-linear model, but hold CFRs constant at 2018 levels between 2019–2030. This reflects our uncertainty in whether past changes in CFRs will continue into the future. Second, we allowed CFRs to change according to the log-linear model between 2000–2030. In Part II of this paper, we established an array of “no-vaccination” scenarios of measles vaccination programs in low- and middle-income countries (LMICs), with different assumptions about the trend of CFR estimates obtained from Part I.

## Methods

### Model overview

We used a previously published log-linear projection model relating CFR to measles incidence, time, and other factors,[5] as shown in the following equation:

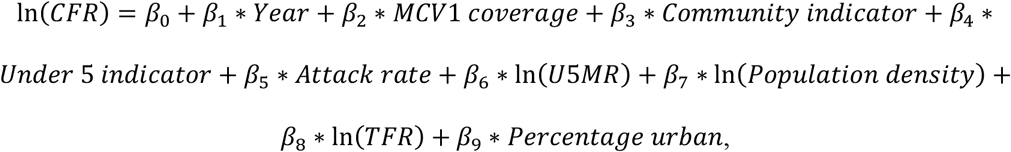

where: *MCV*1 *coverage* = estimated coverage of the routine first dose of measles-containing vaccine (MCV1)[11]; *Community indicator* = an indicator for community-based rather than hospital-based measles; Under 5 indicator = an indicator for children under five years old; *Attack rate* = an approximation of measles attack rate (estimated measles incidence divided by annual birth cohort) [12, 15]; *U*5*MR* = all-cause under-five mortality rate per 1000 live births [12]; *Population density* = population density per square kilometer of land area [12]; *TFR* = total fertility rate [12]; and *Percentage urban* = percentage of population living in urban areas [12].

We stratified the population in 112 LMICs by: (i) under-five mortality rate (</>=50 per 1000 live births); and (ii) world region (Global Burden of Disease regions [16]), and presented all stratifications by age (</>=5 years). The full list of countries by these stratifications is included in Appendix A. For each of these strata, we estimated the measles CFRs across 2000–2030.

The log-linear model was fit to a set of measles CFRs from studies published between 1980 and 2016 [5]. The covariates included 2019 World Bank development indicators [12] and estimates of MCV1 coverage between 2000 to 2018 [11], which allowed us to extend the capability of the model to estimate CFRs up to 2018. The measles incidence estimates required for the model formula were generated from two separate, published measles transmission models used by the Vaccine Impact Modelling Consortium (VIMC) to generate global measles vaccine impact estimates for 2000–2018: the Pennsylvania State University (PSU) model and the DynaMICE model developed at the London School of Hygiene & Tropical Medicine (described in Appendix B and C) [17].

The log-linear model was subsequently used to estimate future CFRs from 2019–2030 in “vaccination” and “no-vaccination” scenarios in the second part of our analyses. We used projected data for covariates, including under-five mortality rate, total fertility rate, percentage of population in urban areas, and population density [18]. Population density was available with annual projections, whereas under-five mortality, total fertility rate, and urban percentage were available by five-year increments [18]. Future MCV1 coverage was projected by the authors from 2018 WHO-UNICEF estimates of coverage [11], assuming a 1% coverage increase per year in line with prior analyses [19]. Future MCV1 coverage was capped at 95%, unless a country had a higher projected coverage as of 2018 in which case the coverage was capped at the maximum coverage reached.

### Part I. CFR scenarios assuming vaccination

We estimated measles CFRs in three scenarios (Table 1). In Scenario-0, we relied on previous estimates of measles CFRs [9], based on a descriptive analysis [10], assumed to be constant across 2000 to 2030. In Scenario-1, we estimated CFRs from 2000 to 2018 using the log-linear model described above with covariates from 2000-2018. This allowed us to examine how these time-variant covariates would affect CFR estimates over time, under the assumption that there is a correlation between CFR, vaccination coverage, measles incidence, and under-five mortality as outlined in previous analysis [5]. In this scenario, the 2018 CFR estimate was assumed to be constant from 2018 to 2030 (held at 2018 levels predicted by the log-linear model), given uncertainty in the trend of covariates due to unknown future data.

**Table 1.** Analytic scenarios.

| **Scenario** | **Model** | **Time-varying**<br>**period** | **Constant period** | **No-vaccination**<br>**scenario** |
| --- | --- | --- | --- | --- |
| Scenario-0 | Constant CFRs [9, 10] | NA | 2000–2030 | (a) Constant CFRs |
| Scenario-1 | Time-varying CFRs [5] | 2000–2018 | 2019–2030 | (a) Constant CFRs<br>(b) Time-varying CFRs |
| Scenario-2 | Time-varying CFRs [5] | 2000–2030 | NA | (a) Constant CFRs<br>(b) Time-varying CFRs |

In Scenario-2, we extended the CFR projection end year from 2018 to 2030, using projected data for covariates, described above. The incidence estimates from 2019 to 2030 were generated from the same measles transmission models used in Scenario-1.

Subsequently, we used the CFR estimates to quantify the impact of estimated CFRs by analytic scenario on estimates of measles deaths from 2000 to 2030, described in Part II, using the PSU and DynaMICE models of measles transmission [9, 20]. We used R statistical software, version 3.6.1, for all analyses [21].

### Part II. CFR scenarios assuming “no-vaccination”

In order to calculate the impact of vaccination, we need to project the burden of disease in the absence of vaccination. Because we had no empirical support for what CFR would be in the absence of vaccination, we tested two alternative approaches. Specifically, we compared the analytic scenarios 1 and 2 in Table 1 to two alternative “no-vaccination” scenarios that assumed: (a) CFRs remain constant; and (b) time-varying CFRs according to the approach used in the comparator “vaccination” scenario. We evaluated how the different assumptions in the “no-vaccination” scenarios affected our estimation on future deaths averted by measles vaccination. These impact estimates were compared to impact estimates where age-specific CFRs in the “no-vaccination scenario” were assumed to be the same as in the corresponding “vaccination” scenario.

## Results

The Scenario-0 CFRs were stratified by age (</>=5 years) for each country; on average across all LMICs, these CFRs were 2.1% for children less than five years of age and 1.0% for children five years of age and older. In comparison, the estimated time-varying CFRs ranged from 3.7% (2.3– 6.3%) in the year 2000 to 1.0% (0.4–3.1%) in the year 2030 for children under five on average across all LMICs, and 1.2% (0.4–3.7%) in 2000 to 0.3% (0.1–1.4%) in 2030 for children five years of age and older. In the year 2018, these estimates were 1.6% (0.7–3.7%) for children under five and 0.5% (0.1–1.9%) for children five and older. The estimated CFRs from 2000 to 2030 by under-five mortality rate and region are listed in Appendix D.

### Impact of constant CFR in “no-vaccination” scenario

Scenario-1 and -2, which assumed the same time-varying CFRs in both the vaccination and no-vaccination scenarios, resulted in fewer deaths averted than Scenario-0 for 112 LMICs; this pattern is consistent for projections from both the PSU and DynaMICE models (Table 2). The percent reductions in measles deaths due to vaccination between years 2000 and 2030 under Scenario-1 (83.3%) and Scenario-2 (83.5%) were both less than the percent reduction in measles deaths under Scenario-0 (85.8%) in the PSU model, and correspondingly estimates for Scenario-1 (91.6%) and Scenario-2 (91.7%) were less than Scenario-0 (92.0%) in the DynaMICE model. Figure 1 displays these results graphically, with the “no-vaccination” upper bound of estimated measles deaths the same across each analytic scenario.

**Table 2.**
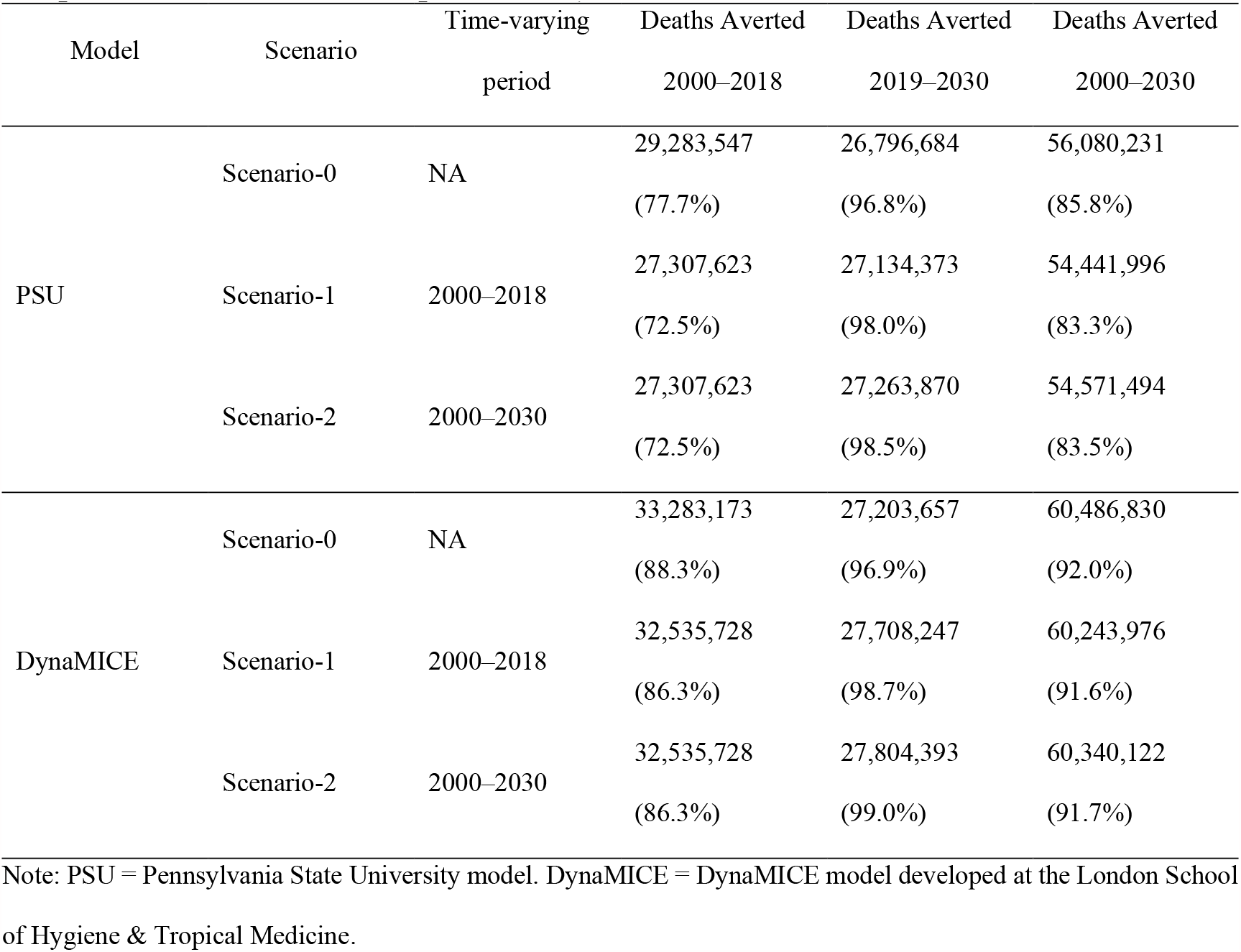
Measles deaths averted due to vaccination for 112 countries across 2000 to 2030, assuming a constant case-fatality ratio in “no-vaccination” scenario (percent reduction compared to no vaccination in parentheses).

**Figure 1.**
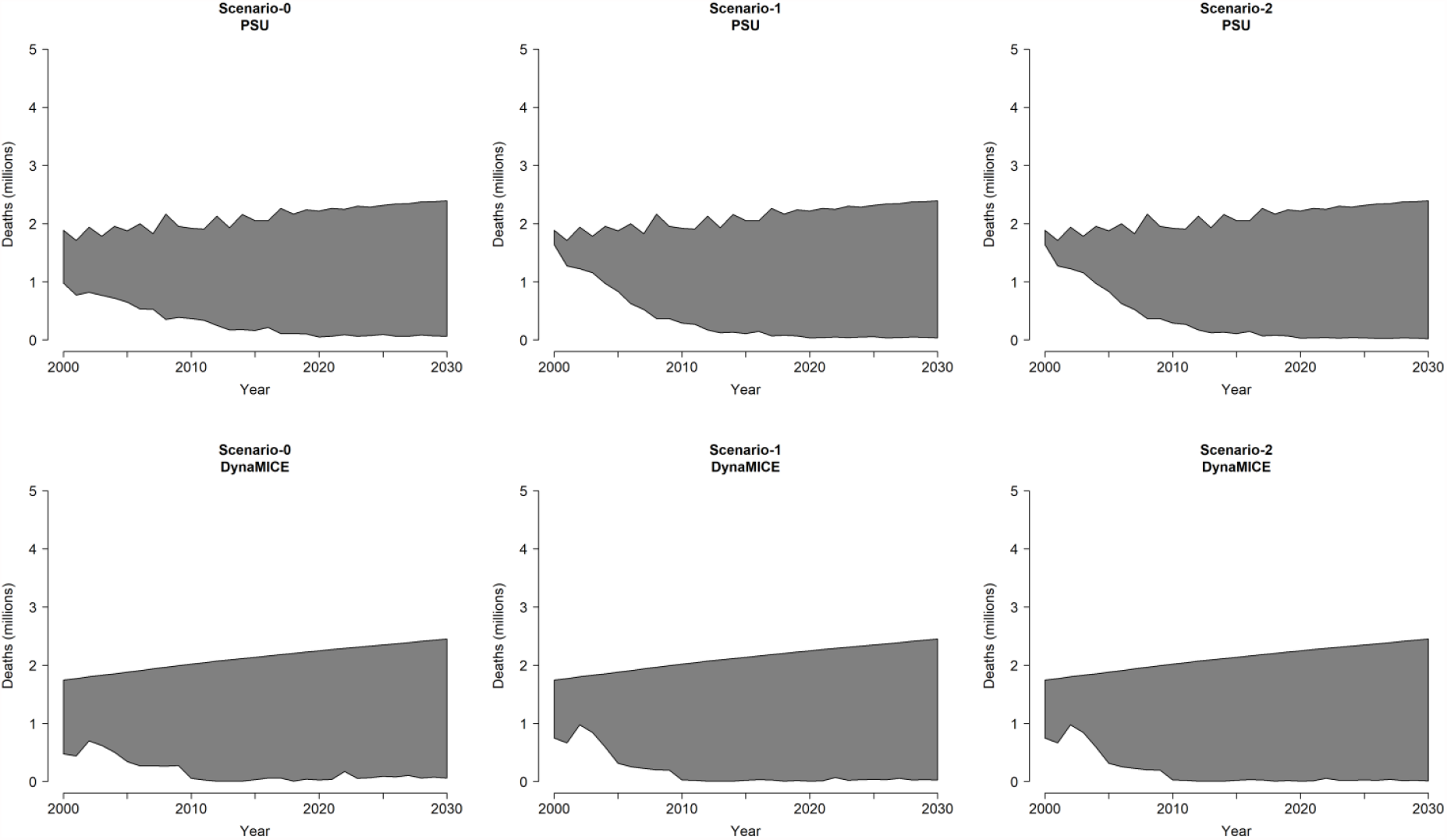
Measles deaths by analytic scenario for 112 countries across 2000 to 2030, assuming a constant case fatality ratio in “no-vaccination” scenario for Pennsylvania State University (PSU) model and DynaMICE model. Note: The top line of each shaded area shows estimated measles deaths in the “no-vaccination” scenario and the bottom line shows estimated measles deaths in the “vaccination” scenario. The shaded region represents the amount of measles deaths averted by vaccination.

### Impact of matching CFR estimation approach in “no-vaccination” scenario

Scenario-0 assumed constant CFRs under both the “vaccination” scenario and the “no-vaccination” scenario. Scenario-1 and Scenario-2 assumed estimated CFRs according to the time-varying, incidence-based methodology for both the “vaccination” scenario and the “no-vaccination” scenario. The percent reduction in measles deaths between 2000 and 2030 under Scenario-1 (86.8%) and Scenario-2 (85.9%) were both greater than the percent reduction in measles deaths under Scenario-0 (85.8%) in the PSU model, and estimates for Scenario-1 (92.4%) were more than Scenario-0 (92.0%) which is more than Scenario-2 (91.9%) in the DynaMICE model (Table 3). Figure 2 displays these results graphically.

**Table 3.**
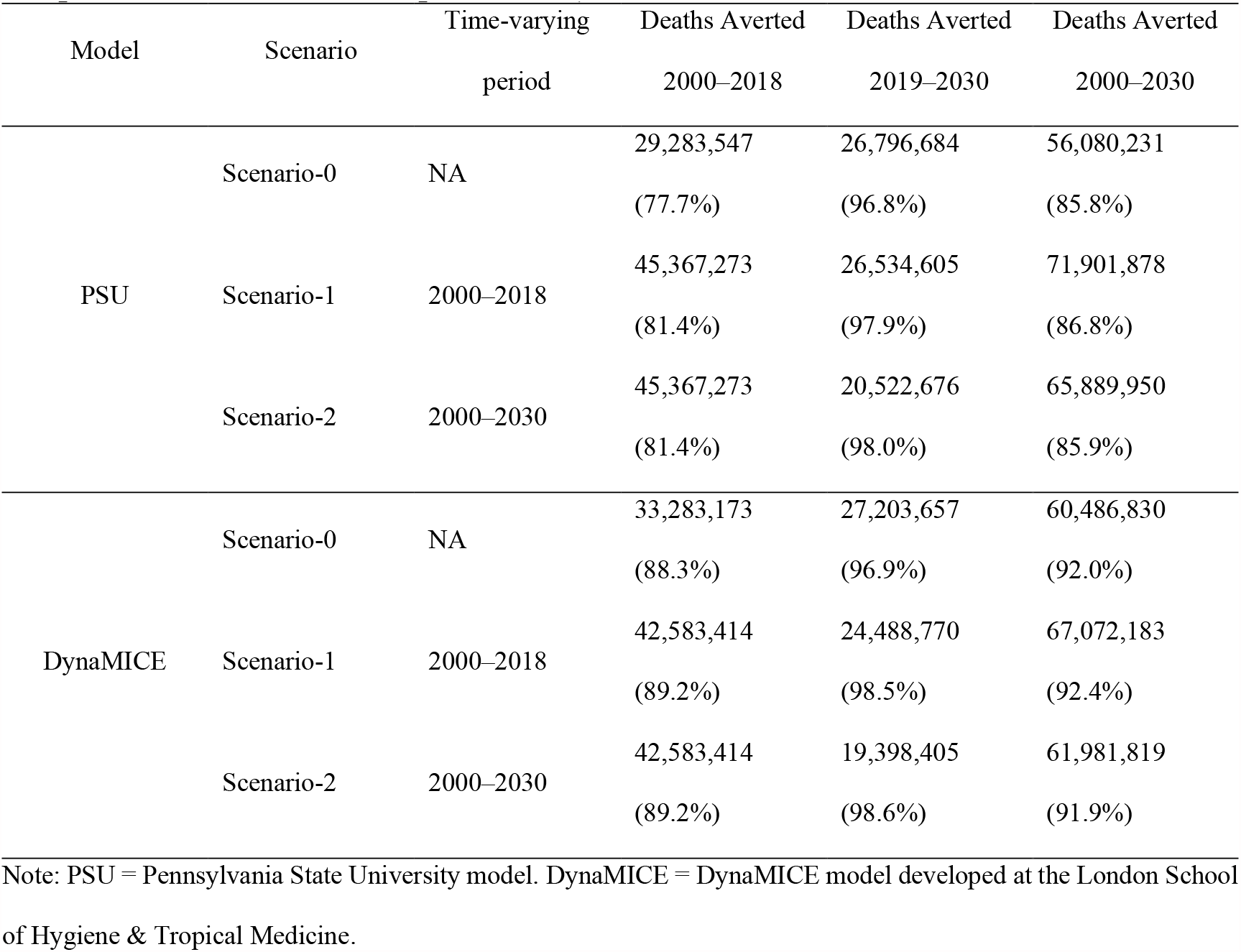
Measles deaths averted due to vaccination for 112 countries across 2000 to 2030, assuming a time-varying case fatality ratio in “no-vaccination” scenario (percent reduction compared to no vaccination in parentheses).

**Figure 2.**
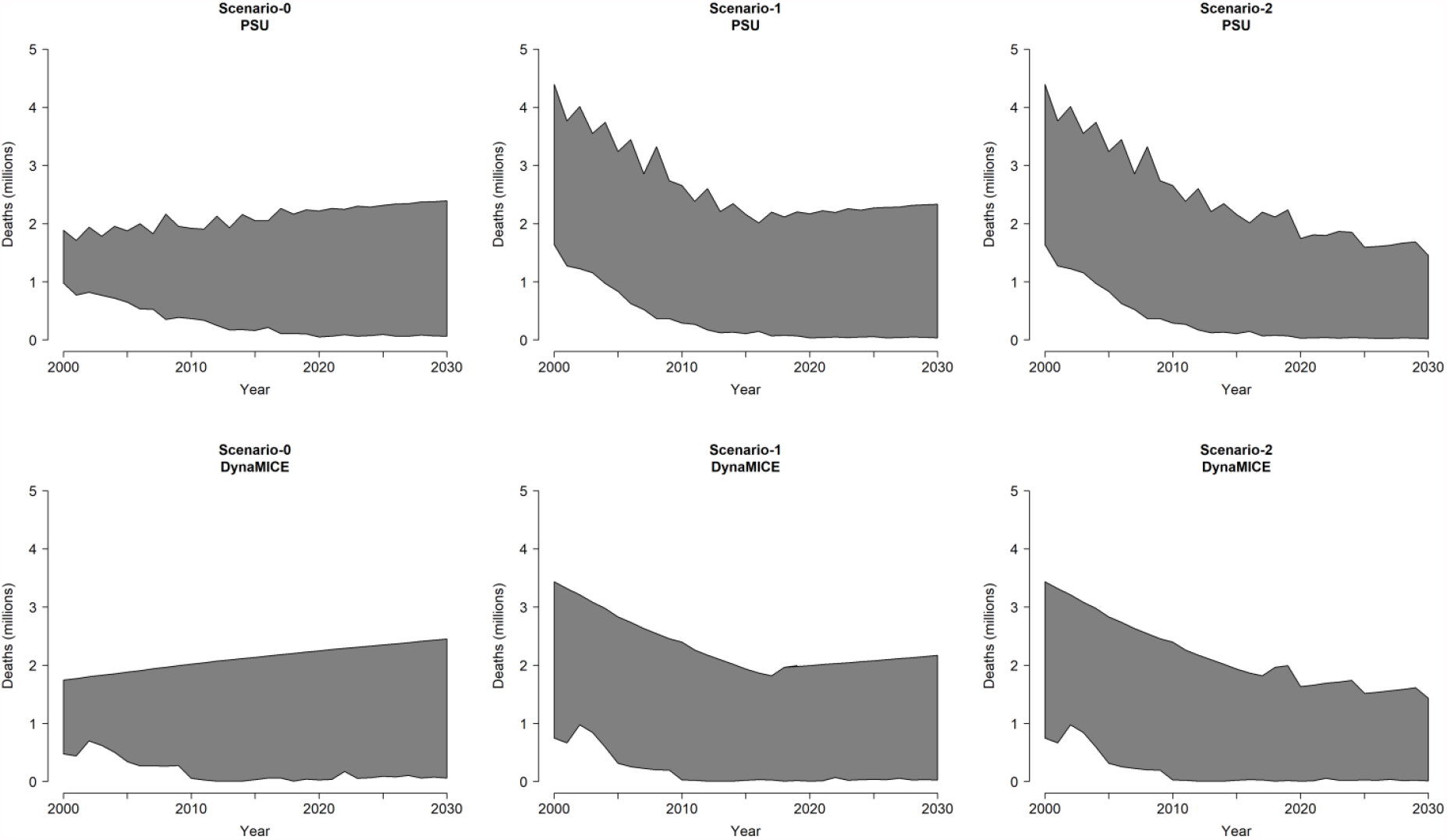
Measles deaths by analytic scenario for 112 countries across 2000 to 2030, assuming a time-varying case fatality ratio in “no-vaccination” scenario for Pennsylvania State University (PSU) model and DynaMICE model. Note: The top line of each shaded area shows estimated measles deaths in the “no-vaccination” scenario and the bottom line shows estimated measles deaths in the “vaccination” scenario. The shaded region represents the amount of measles deaths averted by vaccination.

## Discussion

In this study, we illustrated the effect of important considerations for estimating measles CFRs in the evaluation of measles vaccination impact through alternative “vaccination” and “no-vaccination” scenarios. Our aim was to provide evidence that when measles CFR is dependent on factors such as the incidence of measles and the presence of vaccination, the impact of the vaccine program on mortality risk would depend on these context as well. In order to reflect this context dependence, estimates of measles CFR should be dynamic in time and reactive to differences in scenarios with a transparent methodology that can produce reproducible estimates. Assuming constant measles CFRs produces impact (number of measles cases averted) that grows over time (due to the growth in cases as population grows), but may overestimate the number of deaths that can be averted by measles vaccination in prospective program evaluation and underestimate deaths averted by measles vaccination historically and devalue past gains in retrospective program evaluation. On the other hand, assuming CFRs that decline in the future, in a way that is consistent with empirical observation, leads to impact that is decreasing in the future (because CFRs decline faster than population growth). Recognizing this trend may provide an opportunity to capitalize on these improvements in order to accelerate that decline and create a world in which no (or very few) children die of measles.

It is unclear whether measles CFRs will continue to decrease into the future, particularly given disruptions to vaccination coverage and access to routine health services due to the COVID-19 pandemic. However, the time variant, context dependent approach to estimating measles CFRs can take new covariate data into consideration in subsequent analyses. In this study, we included a scenario to fix future projections of measles CFRs at 2018 levels as an alternative scenario assuming no additional changes to measles CFR from the current context. Specifically, by estimating measles mortality with time variant CFRs from 2000 to 2018 rather than constant CFRs for the vaccination scenario, the percent reduction in measles deaths due to vaccination decreased from 77.7% to 72.5% and 88.3% to 86.3% in the PSU and DynaMICE models, respectively. This reflects higher vaccine coverage in the later part of this time period. However, by aligning both the “vaccination” and “no-vaccination” scenarios with time variant measles CFR estimates from 2000 to 2018, the percent reduction in measles deaths increased from 72.5% to 81.4% and 86.3% to 89.2% in the PSU and DynaMICE models, respectively. The visualizations presented in Figures 1 and 2 highlight that the differences in mortality by year with the dynamic process showed much greater mortality in historical years and lower mortality in future years compared to constant CFRs. In addition, these reductions reflect measles vaccination impact across 112 LMICs, representing a difference of 10–18 million deaths averted. When matching the CFR estimation approach between the “no-vaccination” and “vaccination” scenarios in Figure 2, the rebound effect in estimated measles deaths seen in Scenario-1 holding CFRs constant beyond 2018 is due to increasing population growth, despite the constant CFRs.

There are several common approaches in the literature that account for the uncertainty in CFRs in vaccination program evaluation. To name a few, in a prospective program evaluation study where Nonvignon, *et al*., assessed the impact of rotavirus vaccination in Ghana from 2012 to 2031, the authors used a one-way sensitivity analysis to account for their uncertainty in CFR on their model estimate [22]. In another prospective program evaluation study of pneumococcal conjugate vaccine (PCV13) in India, Krishnamoorthy, *et al*., conducted scenario analyses in which they varied parameter values of mortality, disease event rates, vaccine efficacy, coverage projections, and costs by 10% to evaluate the impact of the program in the most and least favorable scenarios [23]. In a study evaluating the impact of past influenza vaccination in the Netherlands, Backer, *et al*., conducted a probabilistic sensitivity analysis to assess how the uncertainty of their model parameters affected their effect estimates, where CFR was modeled as a normal distribution [24]. Methodological approaches such as these capture the uncertainty in the parameter values of CFR in the model, but they do not automatically account for the potential time dependence of CFR. To account for the time trend in CFRs, health impact models can rely on global burden estimates of disease-specific deaths to derive proportional mortality [25] and statistical models like autoregressive integrated moving average models (ARIMA) have also been used in the vaccine literature [26]. However, in methods that make one-step ahead forecasts such as ARIMA models, long-term forecasting may not be reliable.

In contrast, our study demonstrated a recursive approach where we leveraged a regression model and a transmission dynamic model to account for the time-dependence of CFRs in the estimation of vaccine impact on mortality. Additionally, by using the incidence estimate obtained from a transmission dynamic model as a covariate in our regression model as well as scenario-specific estimates of CFR values in this transmission dynamic model to forecast the impact of vaccination, we were able to capture the long-term dynamic between CFRs, vaccine coverage, and measles incidence in our estimation of vaccine impact on mortality. Further, by explicitly considering the time trend in CFRs in “vaccination” and “no-vaccination” scenarios, we demonstrated that the assumptions made in “no-vaccination” scenarios can affect the estimation of the public health impact of vaccines and other prevention policies on mortality.

There are several limitations to this analysis. First, the functional form for the relationship between the analyzed covariates and measles CFRs is informed by a limited data set of varying quality, as described previously [5]. The Immunization and Vaccine-related Implementation Research Advisory Committee (IVIR-AC) of the WHO in its recent recommendations noted the need for ongoing primary data collection, including “investments in strengthening outbreak investigation and evaluation activities to generate additional primary data” and the “creation of a standard CFR study protocol and a structured data collection tool to improve comparability of studies” [27]. Second, the log-linear model to predict measles CFR does not necessarily represent causal relationships between the covariates of interest and the outcome. Additional factors that are important considerations for mortality risk, such as malnutrition and treatment status of the individual cases, would require future data collection efforts to supplement currently available data on measles CFRs [5]. We addressed these limitations by estimating two different versions of the “no-vaccination” scenario. While the causal effect of variables not included in the model might be captured in the variables we included, it is unclear which variables should apply to the “no-vaccination” case. Third, there may be additional uncertainty in the impact estimates according to the assumptions of each measles transmission model, described previously [9, 20, 28, 29].

In order to better estimate the impact of public health interventions, this study can shed light on the effects of alternative assumptions to project future scenarios evaluating intervention impact and provide guidance for developing appropriate “no-vaccination” scenarios. We would expect to see similar relationships between disease management and health system capacity with the risks of disease outcome over time. For example, hospital-fatality risks for COVID-19 tend to increase as availability of hospital beds and ventilators decreases; on the other hand, there are temporal changes in fatality risk due to improvements in treatment. To assess the consequences of public health interventions, impact estimates should consider the effect of “no-vaccination” scenario assumptions on health impact model parameters such as measles CFR, in order to serve the goals of both: estimating the historical and current impact of interventions; and projecting estimated impact for alternative scenarios according to intervention strategies and investment decisions. As additional and improved empirical evidence of program implementation becomes available to inform predictive models, we can continue to improve predictions and uncertainty of the effect of public health interventions, such as measles vaccination.

## Data Availability

The data underlying this article will be shared on reasonable request to the corresponding author.

## Acknowledgements

The authors thank Matthew Hanson and Stéphane Verguet for their collaboration and support on the previous analysis that this work builds on.

## Presentations of work

Preliminary results of this work were presented at a meeting of the Immunization and Vaccines-related Implementation Research Advisory Committee of the World Health Organization (March 2021).

## Appendix A. Country list by under-five mortality rate (U5MR) and region

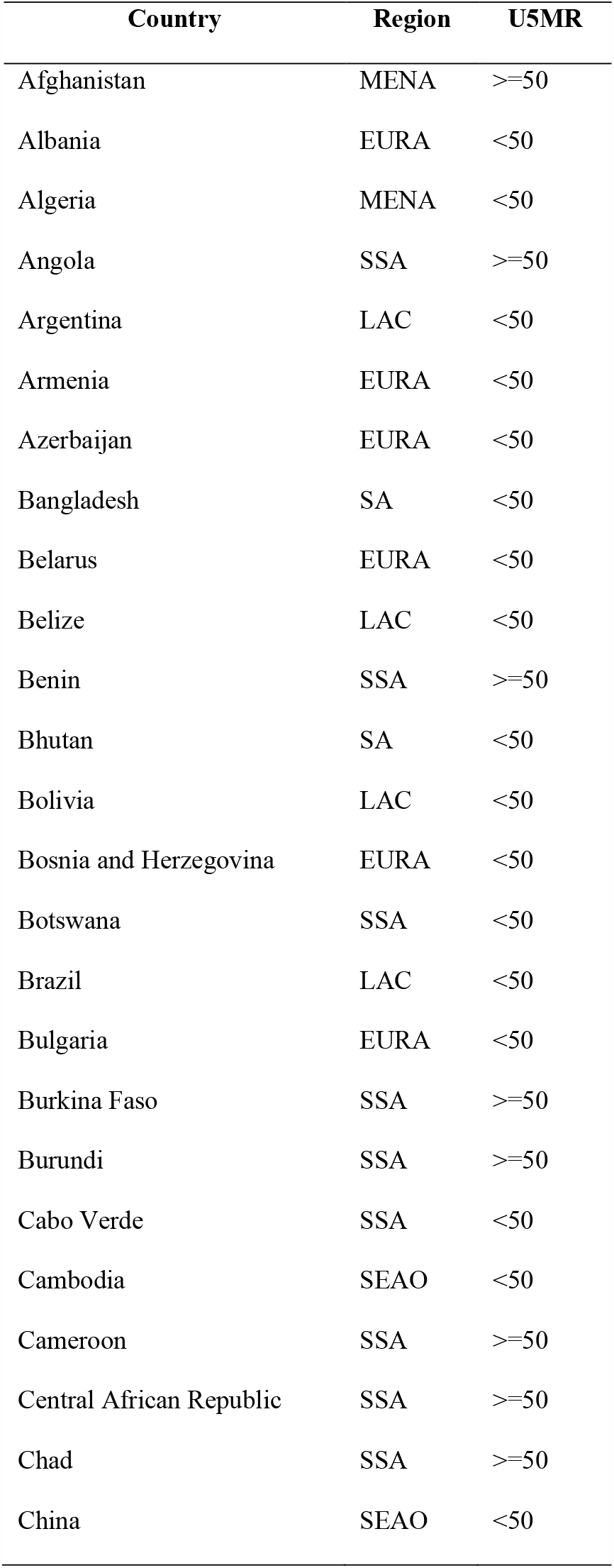
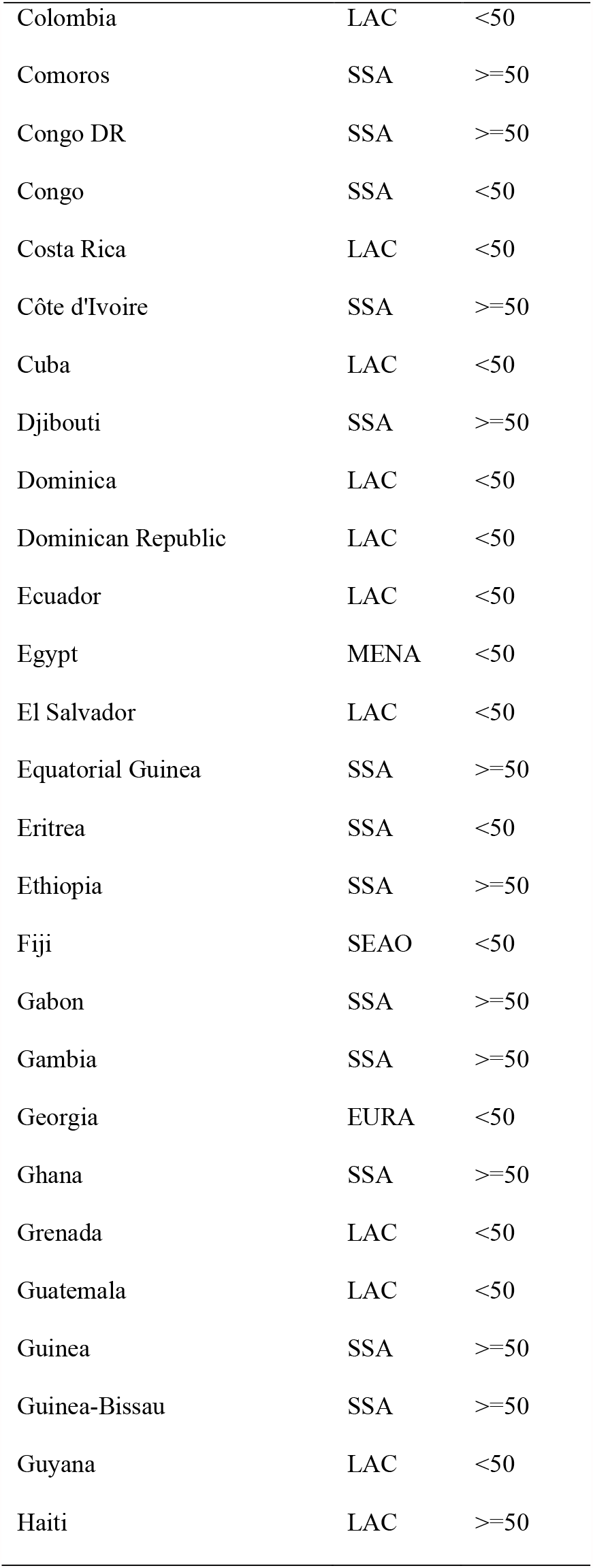
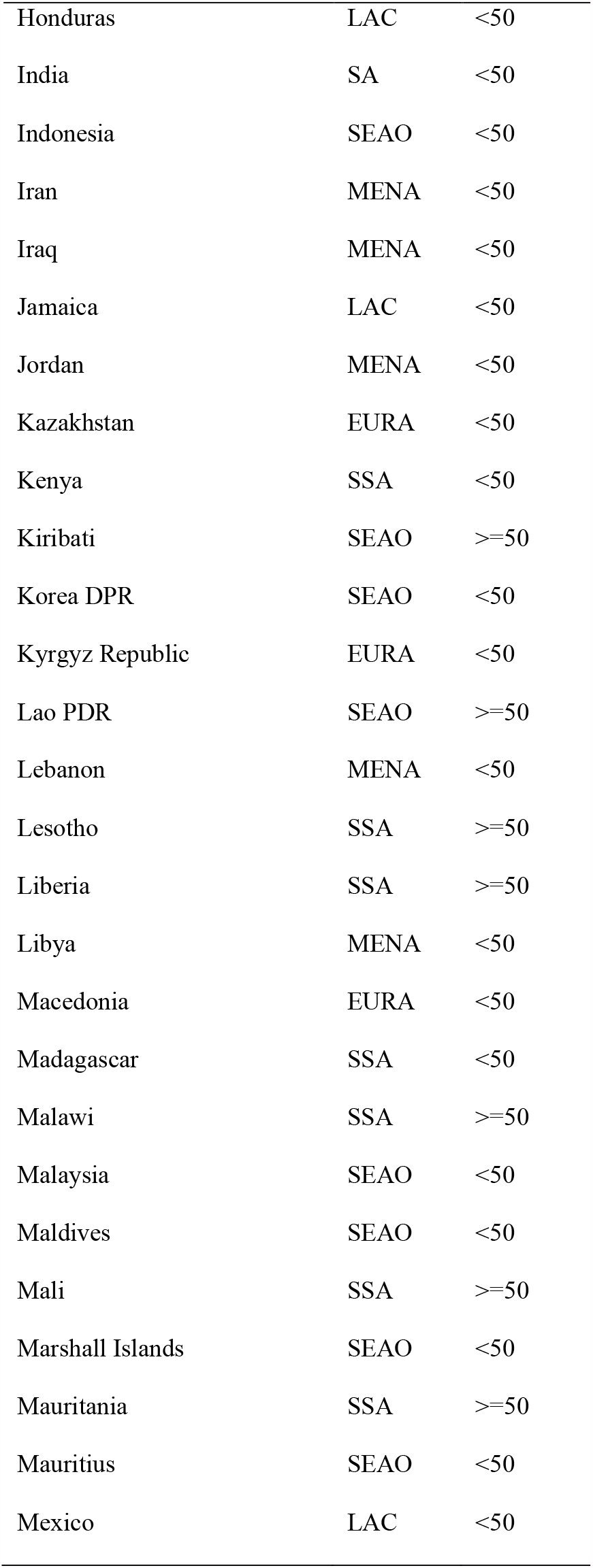
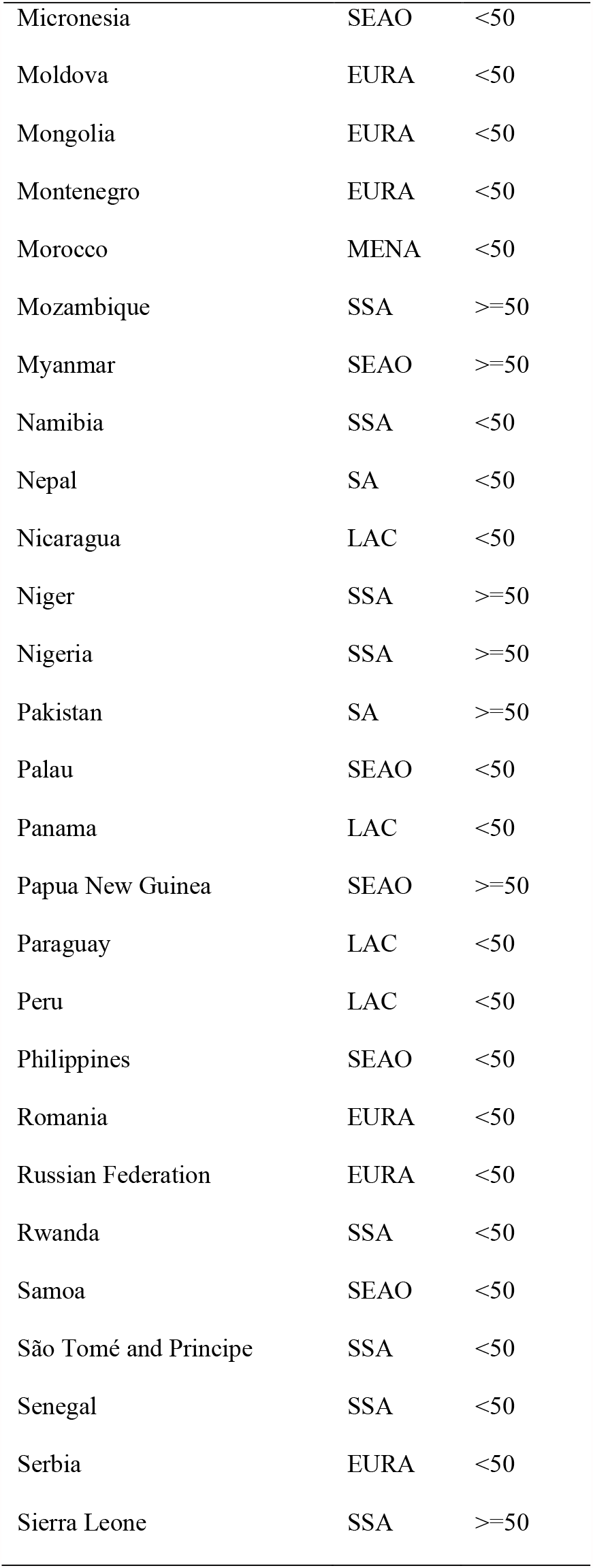
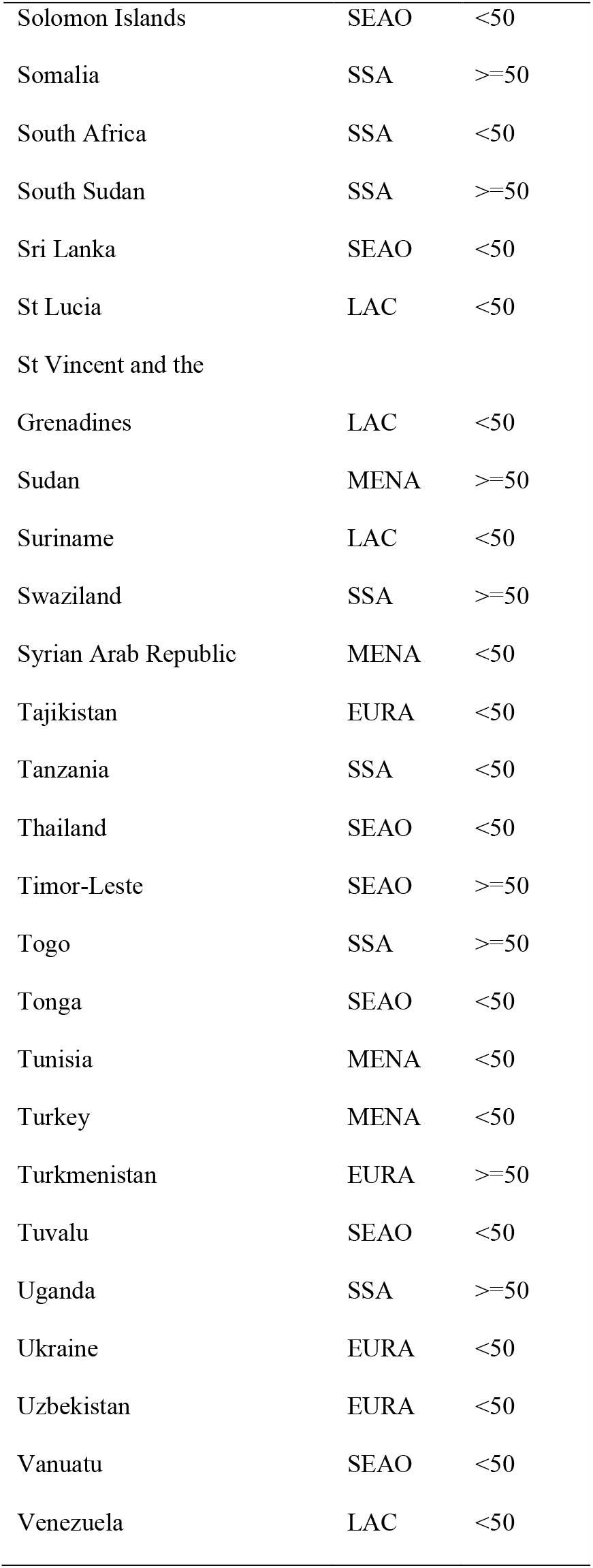
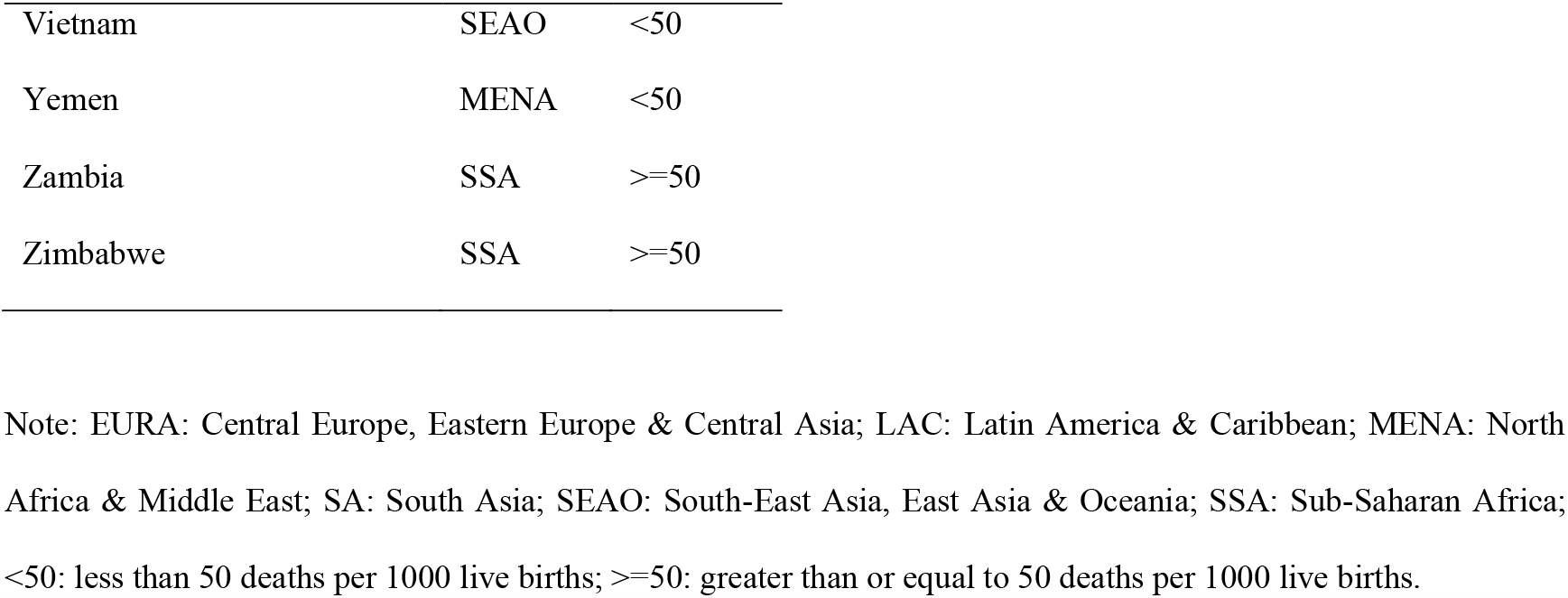

## Appendix b. DynaMICE Model Overview^1^

The Dynamic Measles Immunization Calculation Engine (DynaMICE) is a dynamic age-stratified population model of measles transmission dynamics to estimate the public health impact and cost-effectiveness of routine vaccination programs and supplementary immunization activities (SIAs) in low and middle-income countries.^2^ It was originally developed for work with the World Health Organization and received inputs from investigators at Harvard University and Montreal University as well as LSHTM. It was subsequently used to inform vaccine impact projections for Gavi, the Vaccine Alliance and the Bill & Melinda Gates Foundation. The model provides a flexible framework that can be adapted to different countries with the aim to study several vaccination scenarios based on available data sources. For example, the model has been adapted to characterize measles transmission and dynamics in India, based on measles data from the Million Deaths Study,^3^ as well as to quantify the impact of adding interventions to measles SIAs in India by interfacing with the Lives Saved Tool (LiST).^4^

As measles is a highly transmissible childhood infection, disease dynamics are inextricably linked to population structure and demographic parameters. To enable precision in the estimation of disease burden and the contact processes that drive transmission, the model is age-stratified to include weekly age groups from birth to 3 years of age, and yearly age-groups from 3-100 years of age. The underlying epidemic model is a compartmental SIR model, where individuals can either be susceptible (S) to measles, infected (I) or recovered (R) with life-long immunity. After a certain duration of maternal immunity, births replenish the pool of susceptibles that in the absence of vaccination fuel periodic outbreaks of measles driven by the magnitude of the birth rate and the strength of seasonality in transmission parameters. Susceptibles get infected through contact with infected individuals, with mixing determined by age-dependent contact patterns. The contact matrix used for this exercise is the POLYMOD contact matrix for Great Britain^5^ as it was best able to reproduce transmission dynamics across a range of countries, but this can be updated to represent local population age structure. Following infection, individuals either recover and gain lifelong immunity, or die as described by country-specific age-dependent case fatality ratios (CFRs).

Routine vaccination is modelled through first- and second-dose measles-containing vaccine (MCV1 and MCV2) schedules (corrected for the right cohorts) with the additional option of including SIAs. Vaccines are assumed to be “all or nothing” with effectiveness equal to 84% for the first dose among children under the age of one year, 93% for the first dose among children over the age of one year, and 99% for both doses,^6^ with life-long vaccine-induced immunity.

## Appendix C. PSU Model Overview^7^

The PSU measles model is a semi-mechanistic, age-structured, discrete time-step, annual SIR model. Unlike conventional SIR models, which describe dynamics at the scale of an infectious generation (TSIR REF)^8^ or finer (basic REF)^9^, it models the aggregate number of cases over one-year time steps. While this is coarse relative to the time scale of measles transmission, it matches the annual reporting of measles cases available for all countries since approximately 1980 for all countries through the WHO Joint Reporting Form (JRF). To account for the fine-scale dynamics that are being summed over a full year, the model describes the number of infections (I_i,t_) in country *i* and year *t*, and age class *a* as an increasing function of the fraction, p_i,t_, of the population susceptible in age class *a* at the start of year *t*, S_i,t_:

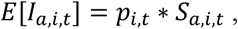

where *E* [·] indicates the expectation and, *p*_*i,t*_ is a country and year specific annualized attack rate modeled as:

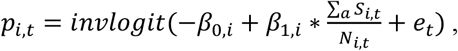

where invlogit() indicates the inverse logit function, N_i,t_ is the total population size in country *i* and year *t* over all age classes, and e_t_ is a Gaussian random variable with mean 0 and variance σ ^2^.

The parameters *β*_0,*i*_, *β*_1,*i*_, and σ^2^ are fit to each country independently using a state-space model fitted to observed annual cases reported through the JRF from 1980-2016 as described by Eilertson, *et al*.^10^ Historical population and vaccination coverage values are provided by WHO as described by Simons, *et al*.^11^

The number of susceptible individuals in each single-year age class *a* (*a*=2,…, 25) is equal to the number not infected in the previous year, nor immunized through supplemental immunization activities (SIAs). The number susceptible is further deprecated by the crude death rate. The efficacy of doses administered through SIAs is assumed to be 99%. The number of susceptible individuals in age class *a*=1 is assumed to be 50% of the annual live birth cohort; this assumes that all children have protective maternal immunity until 6 months of age. Age class *a*=2 and *a*=m is assumed to receive a first and second dose (respectively) of routine measles vaccination before the start of the time step; thus, the number susceptible is further reduced by the product of the coverage and the efficacy. Efficacy is assumed to be 85% and 93% for the first dose in countries delivering at 9m and 12m of age, respectively, and assumed to be 99% for the second dose.

Deaths are calculated by applying an age- and country-specific case fatality ratio (CFR) to each country. CFRs for cases below 59 months of age for all countries were taken from Wolfson, *et al*.^12^; CFR for cases above 59 months of age are assumed to be 50% lower than those applying to under-5s.

Forward simulations of this model assume random variation in the annual attack rate according to the parameter σ^2^. Further, each forward simulation draws *β*_0,*i*_, *β*_1,*i*_ at random from the joint 95% interval estimate of each parameter. Future vaccination coverage values, for routine and SIAs, are assumed known and future birth and death rates are assumed known.

## Appendix D. Estimated measles case fatality ratios (CFRs) from 2000 to 2030 by model, vaccination scenario, age, under-five mortality rate (U5MR), and region

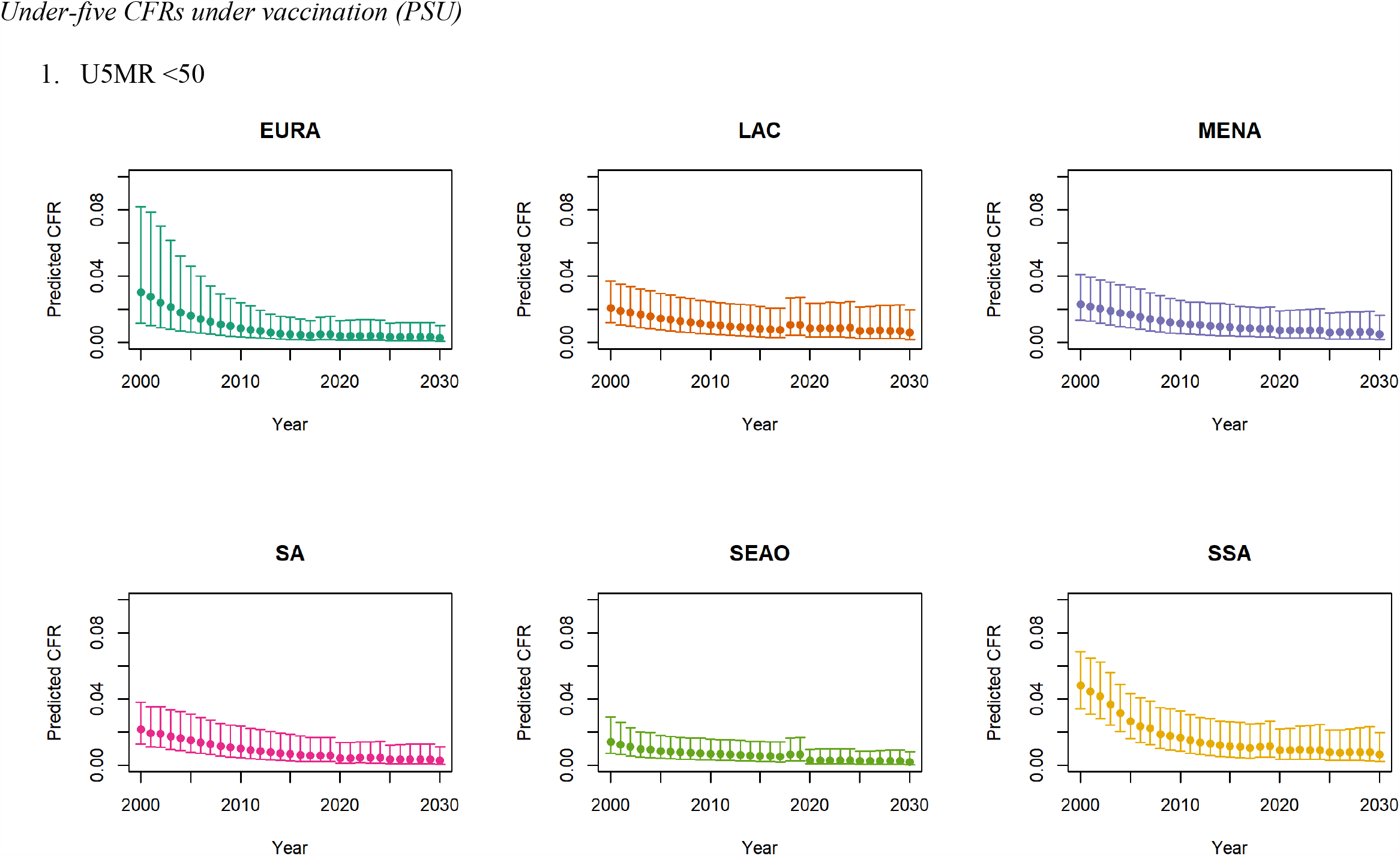
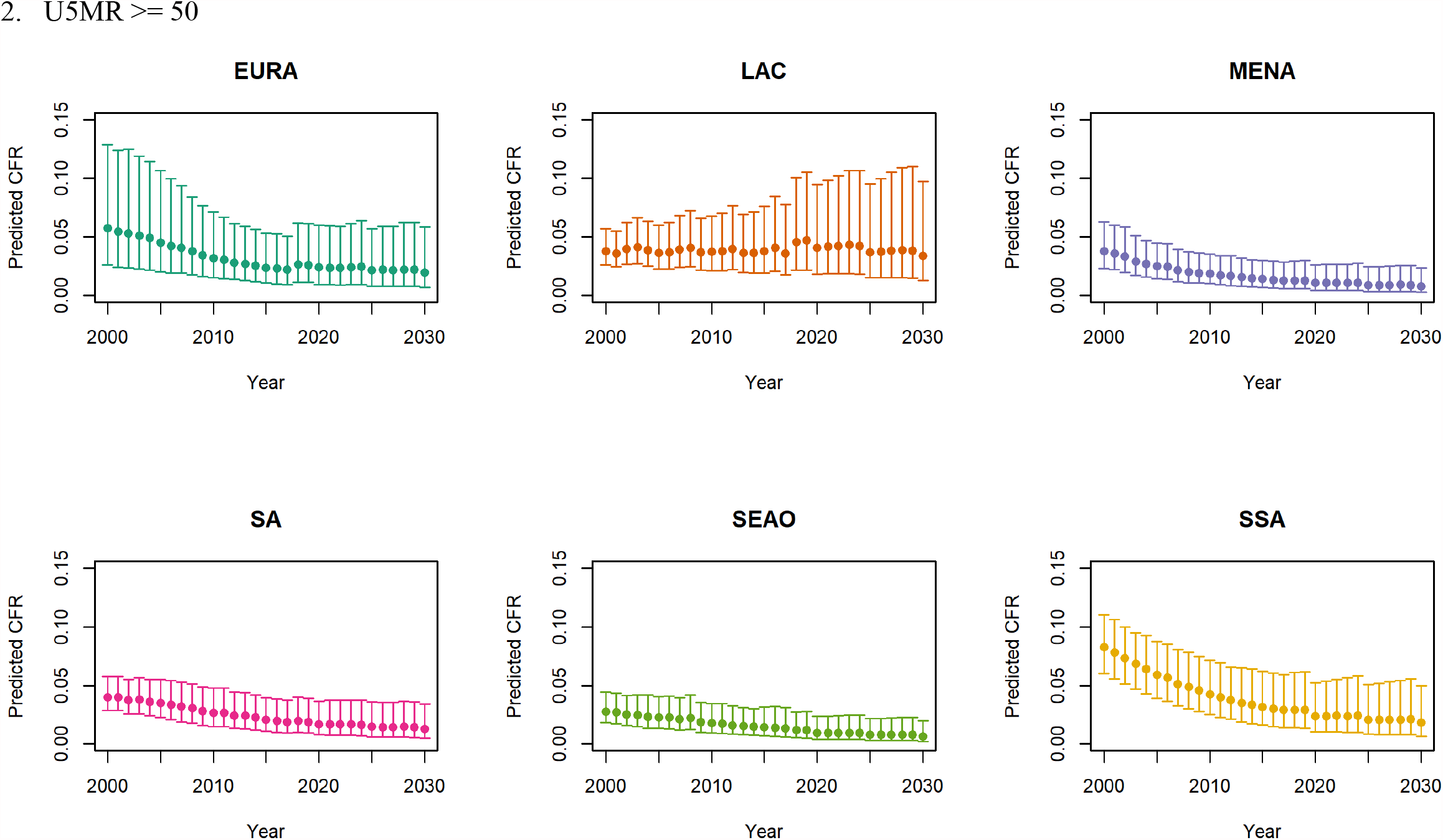
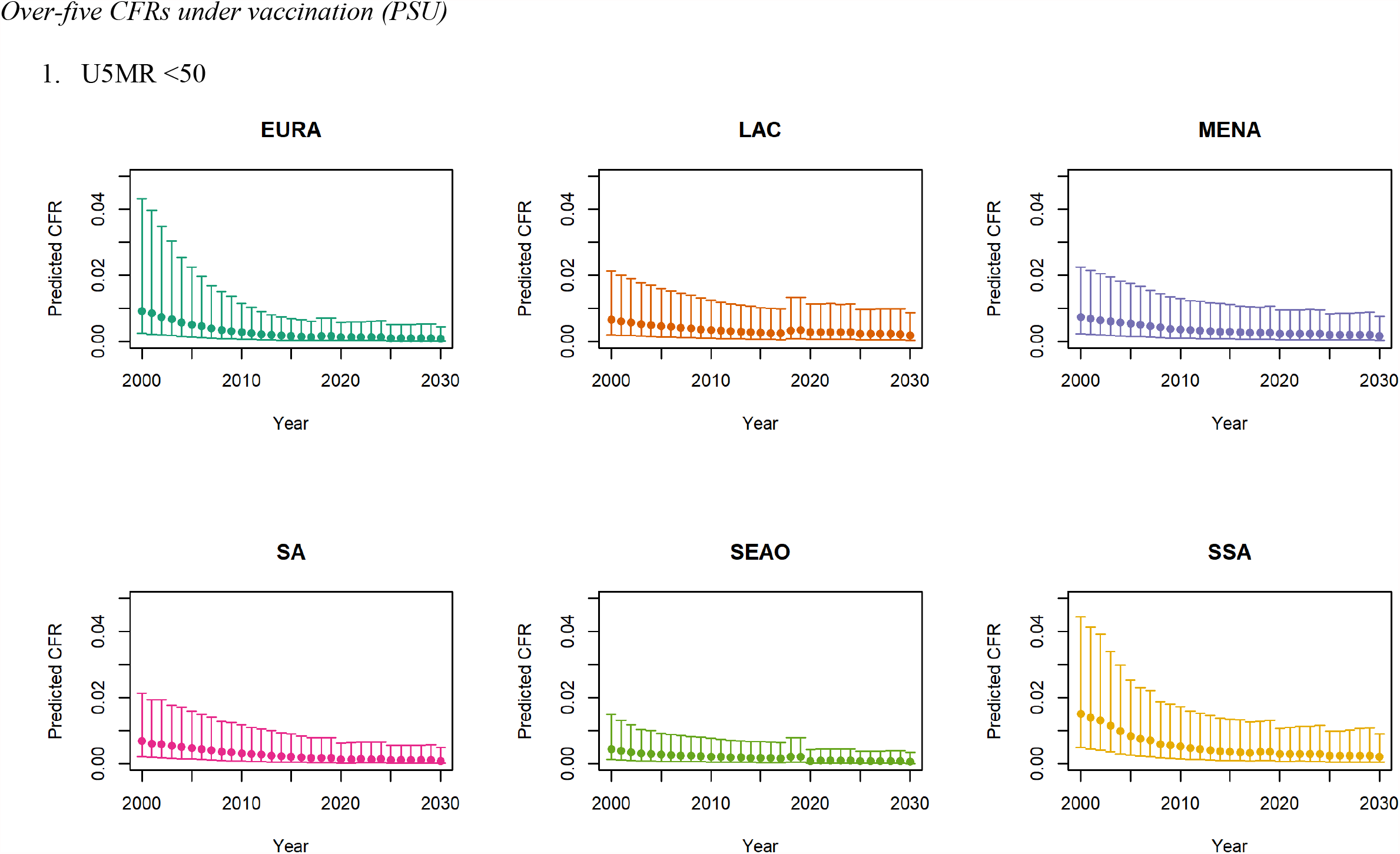
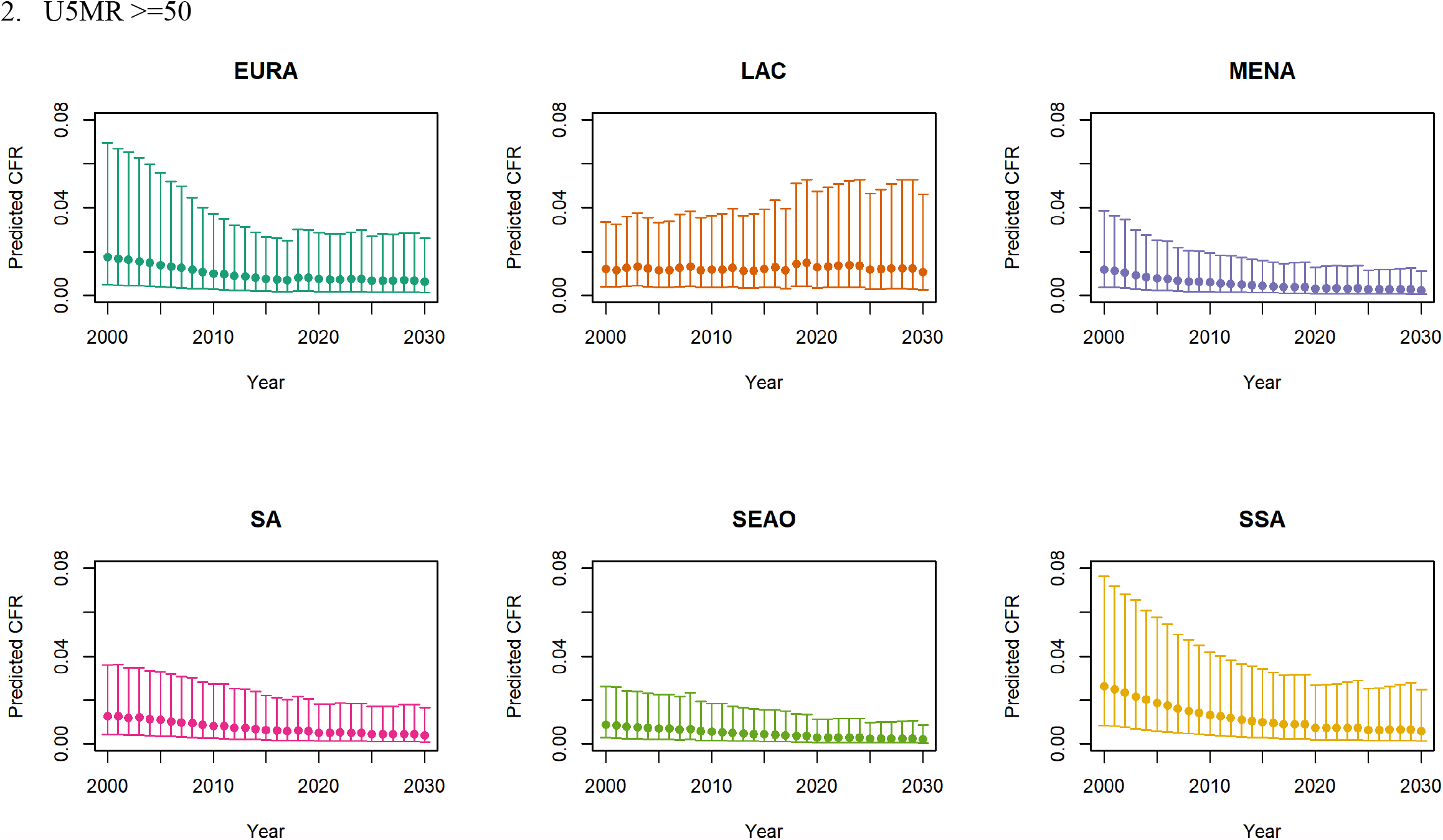
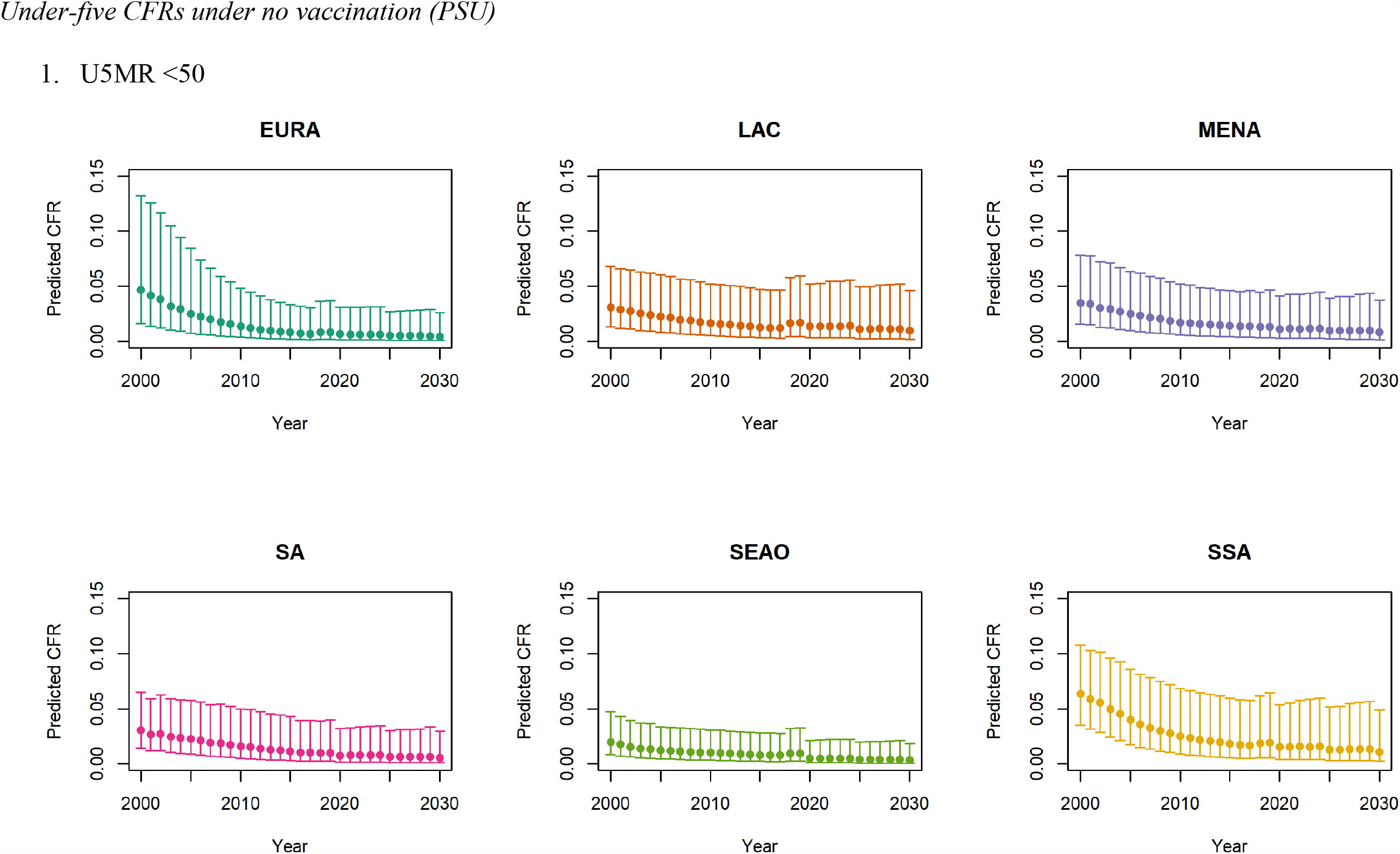
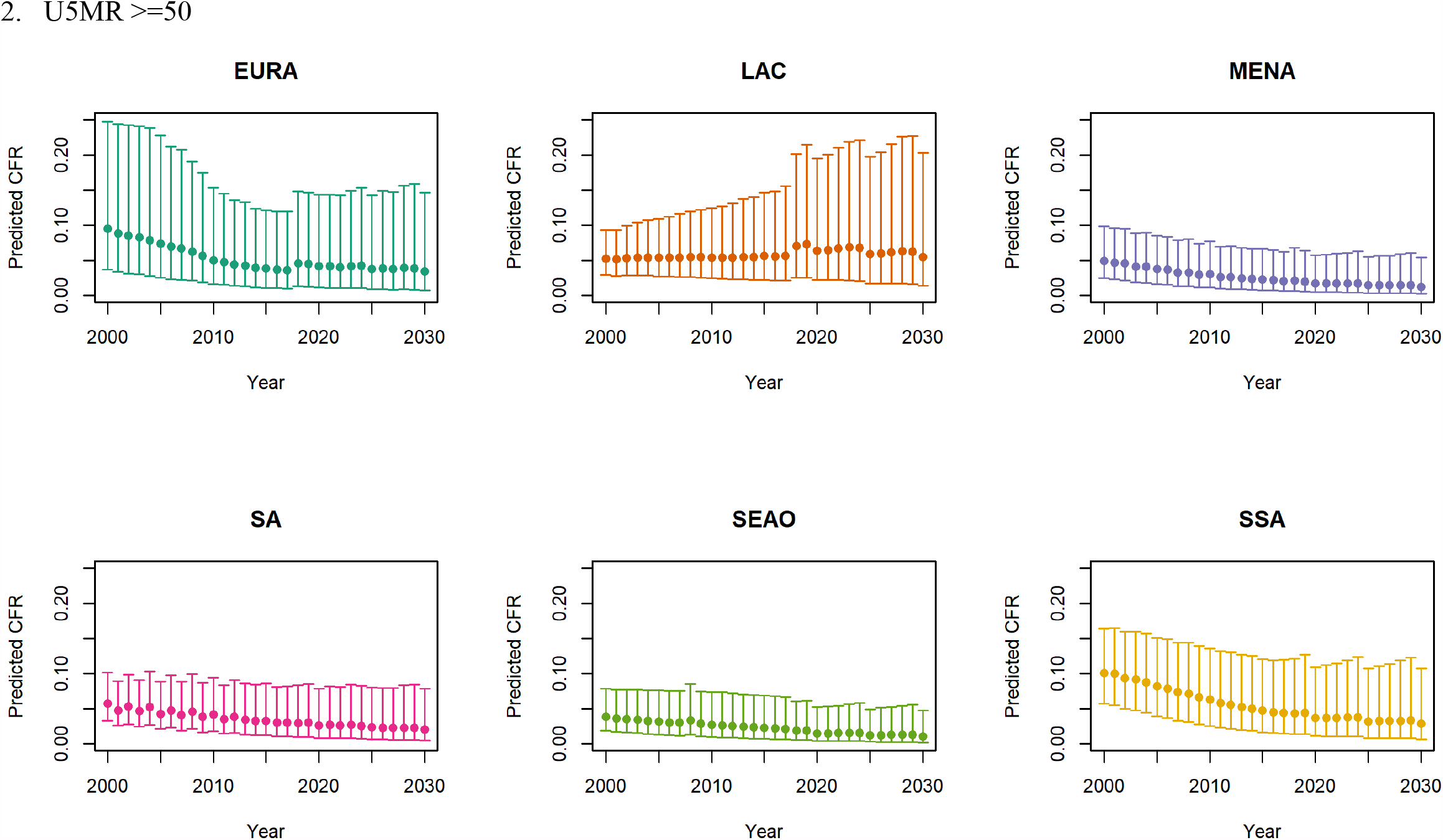
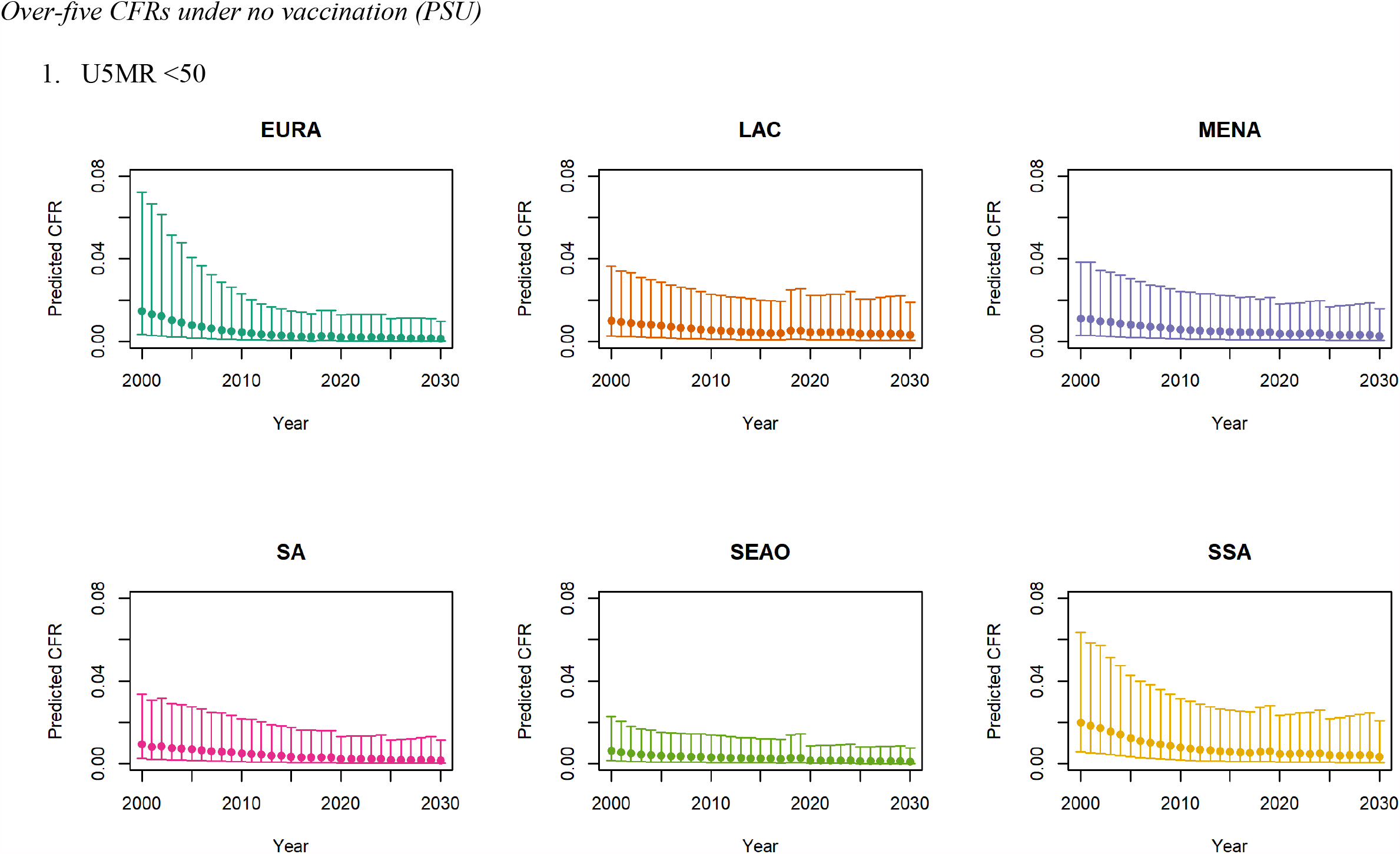
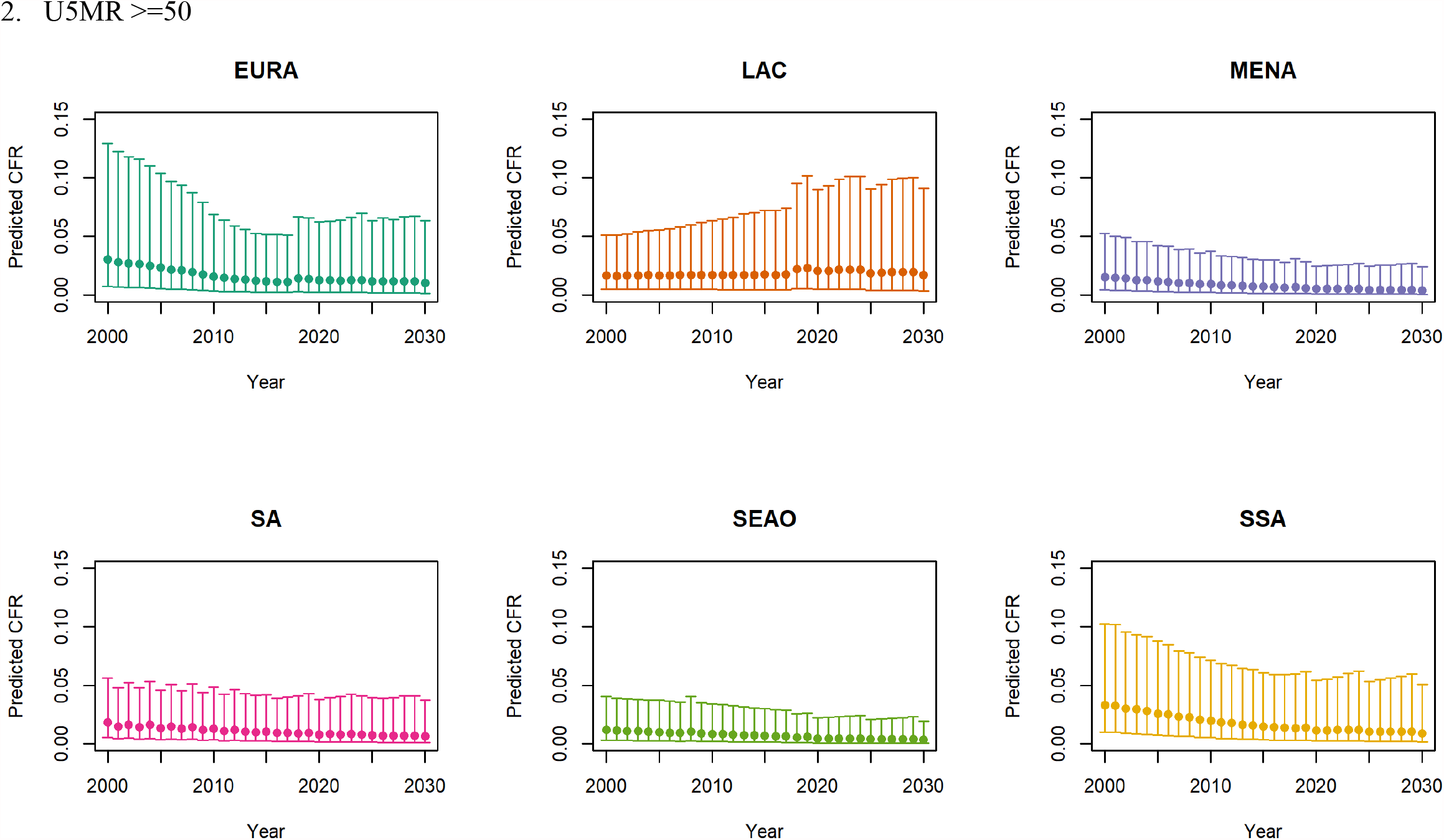
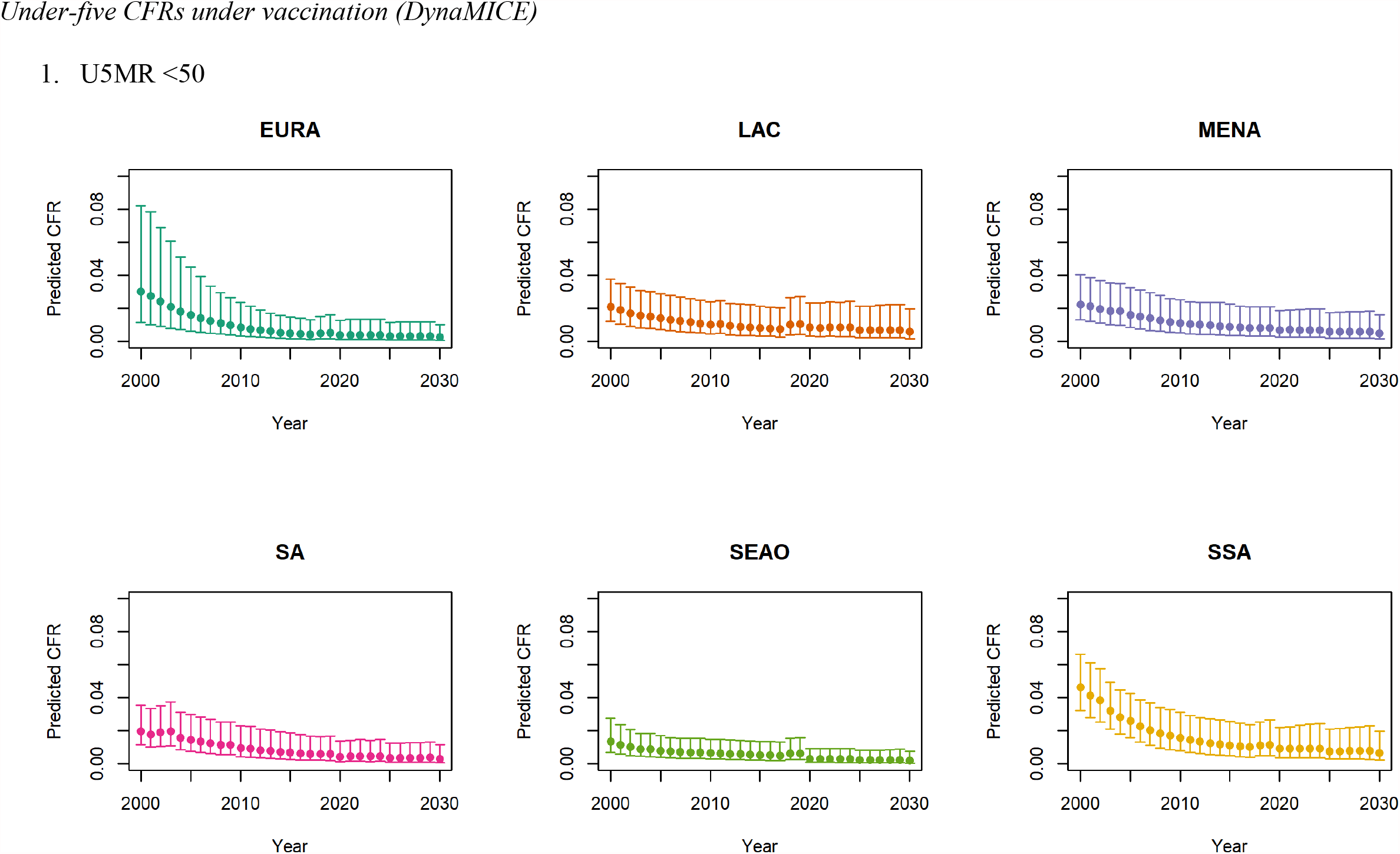
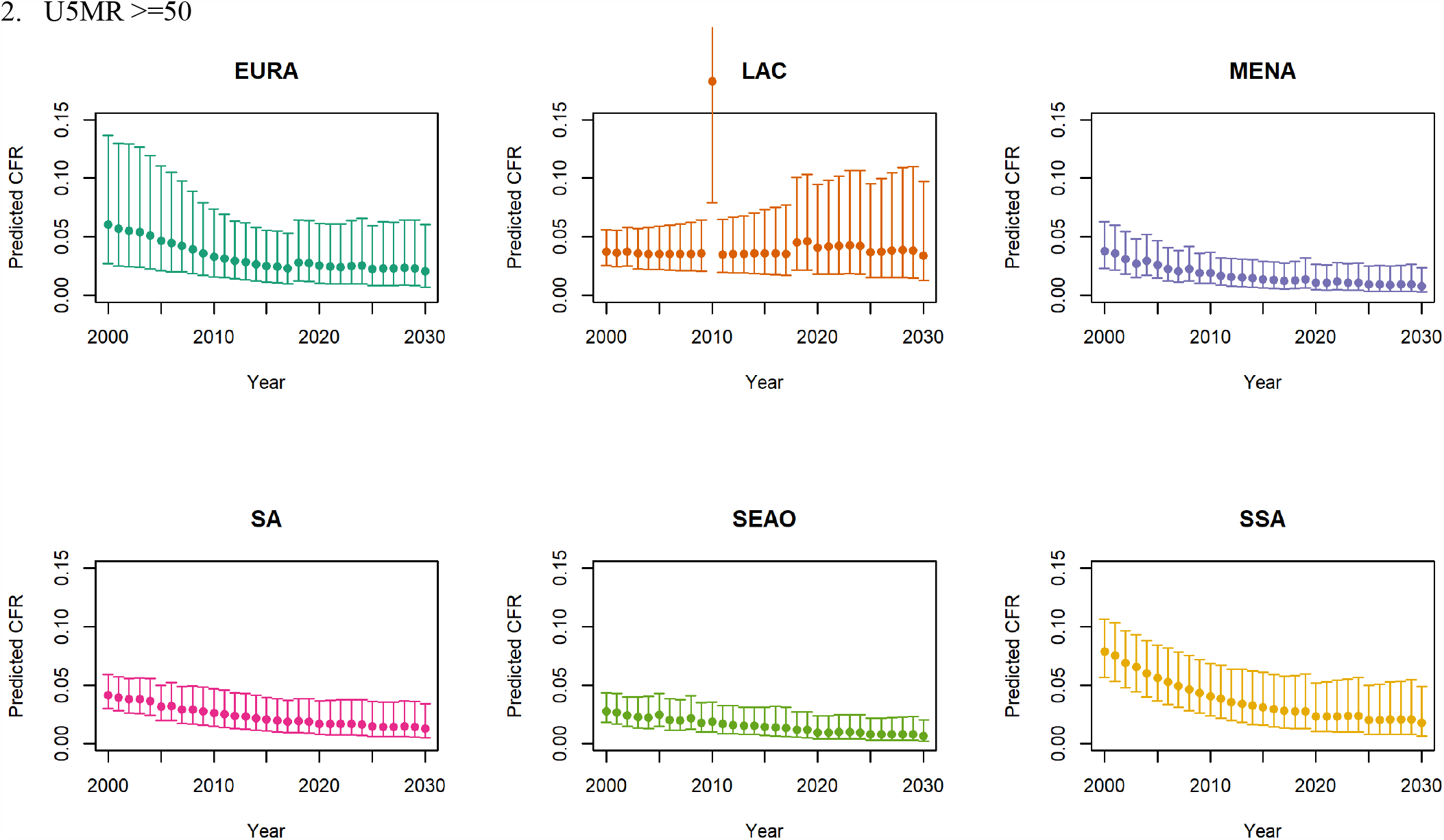

Note: The LAC region at >=50 U5MR only includes the country of Haiti, which experienced a significant shock to covariates due to the 2010 earthquake.

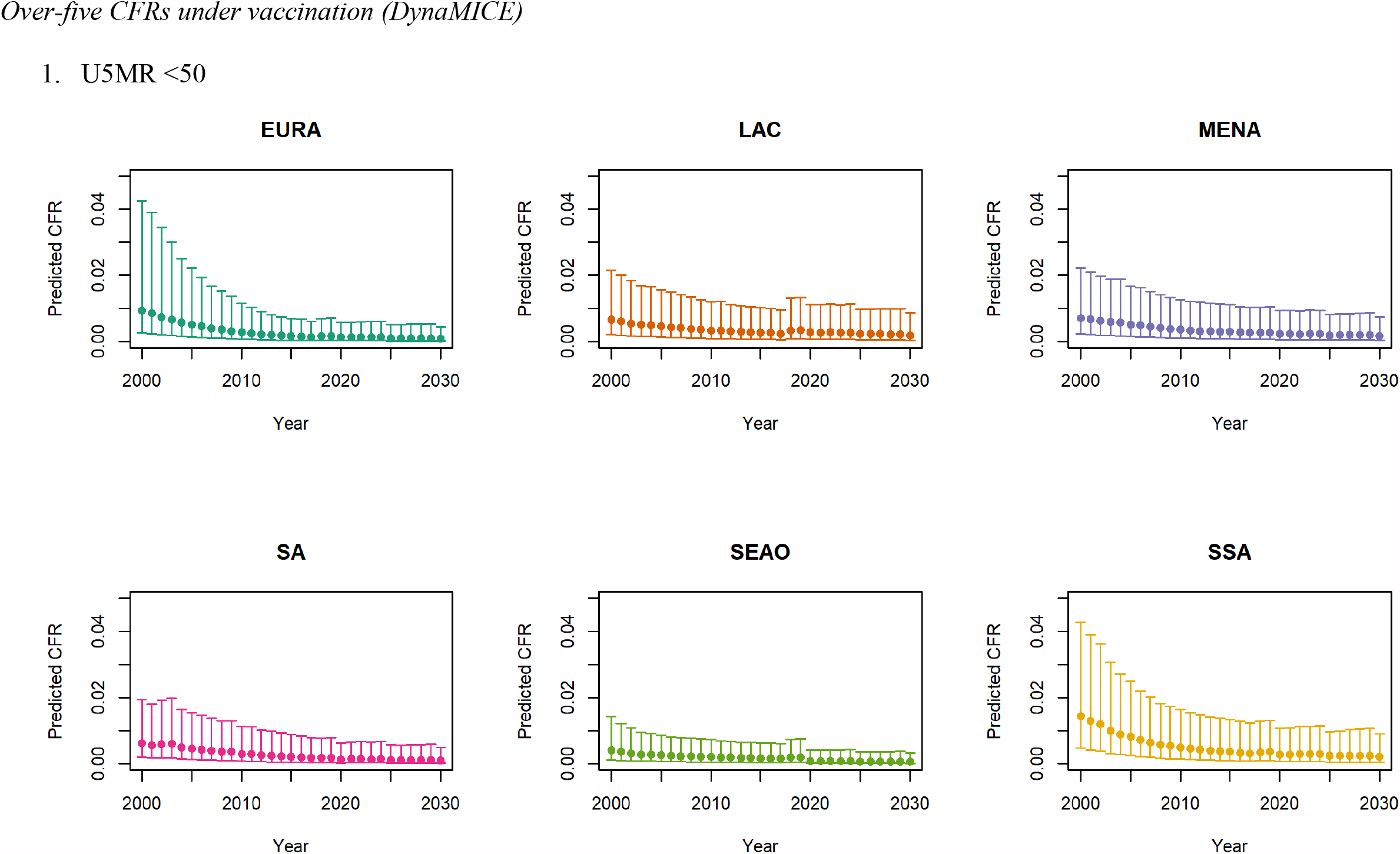
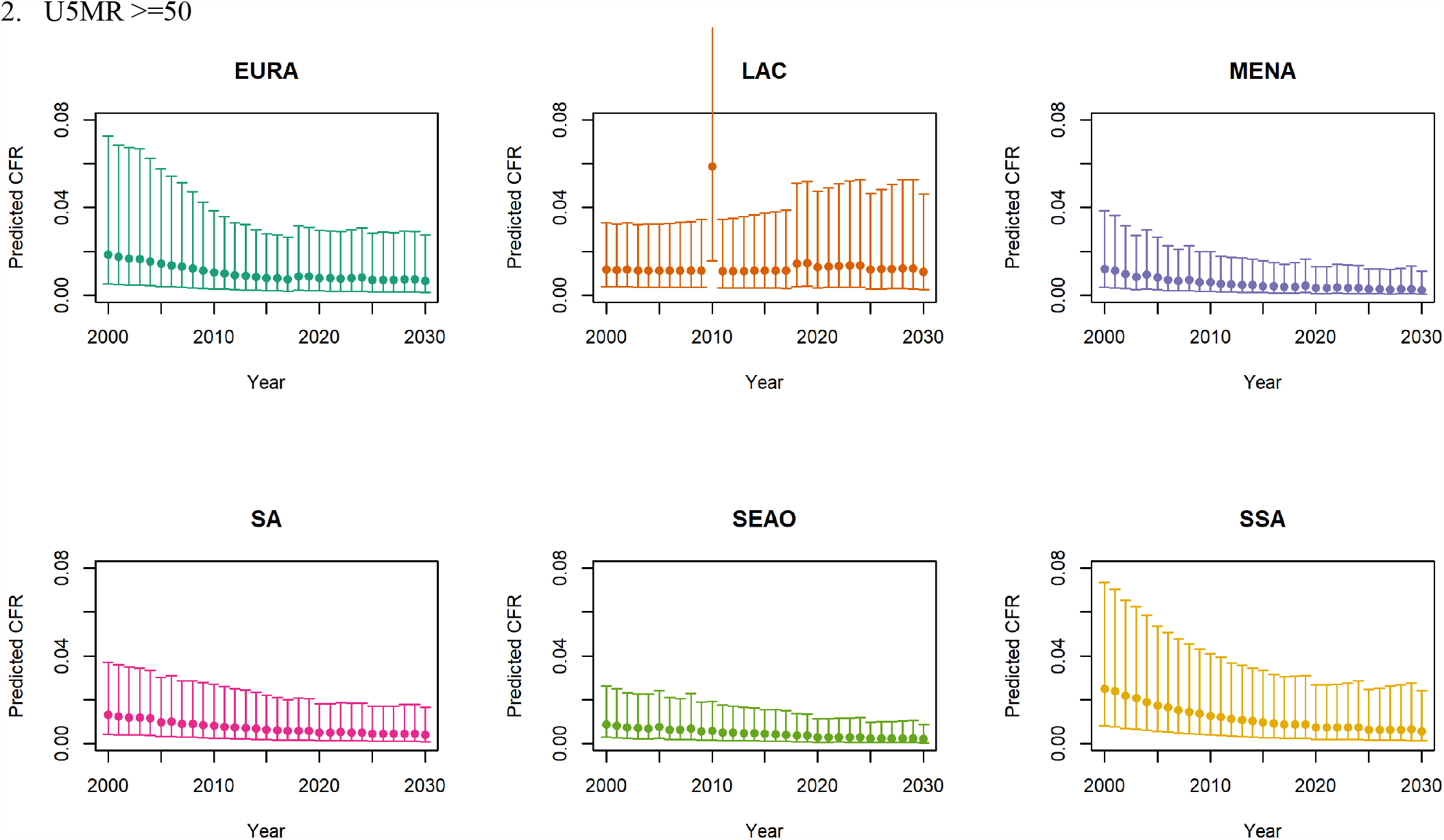

Note: The LAC region at >=50 U5MR only includes the country of Haiti, which experienced a significant shock to covariates due to the 2010 earthquake.

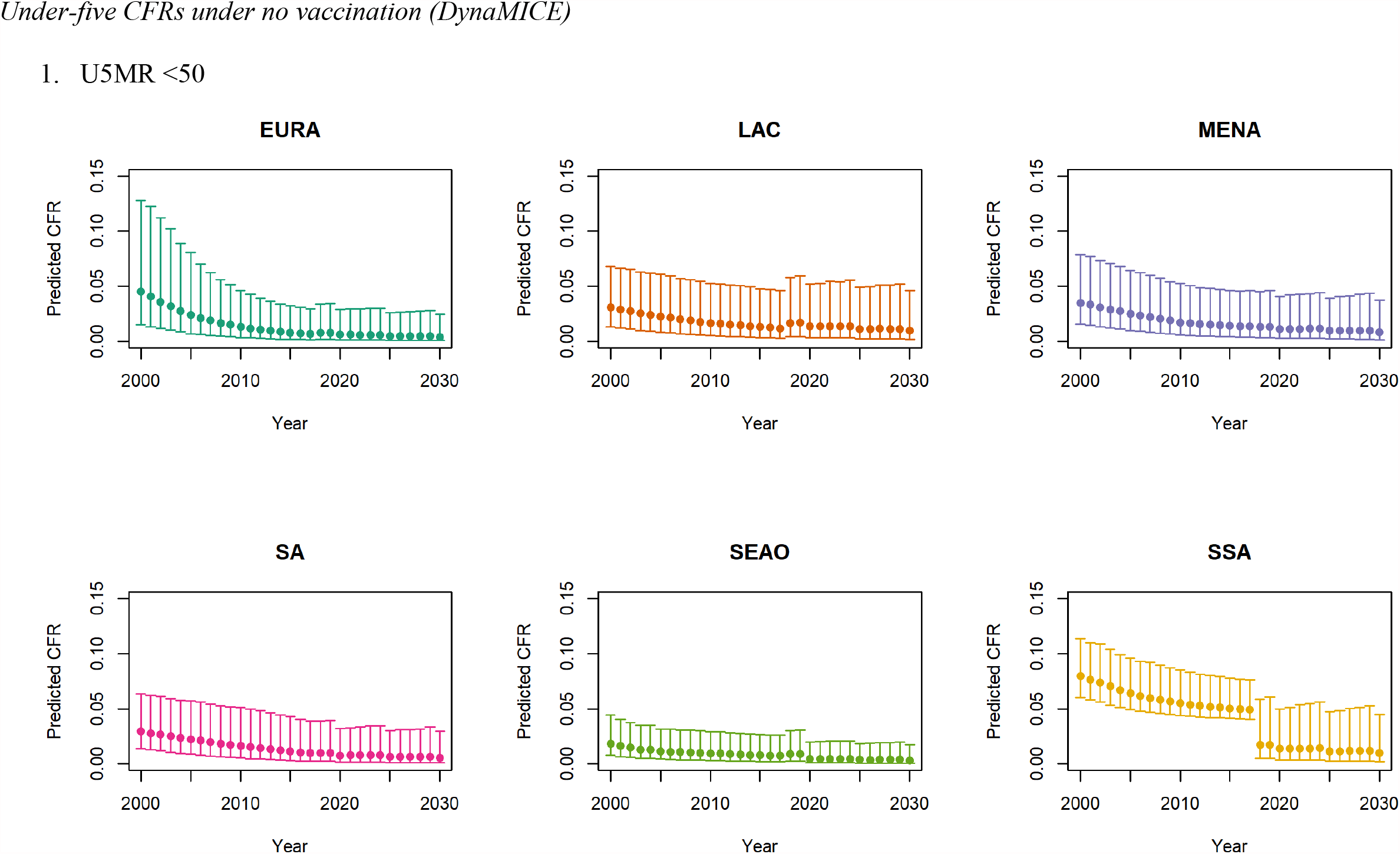
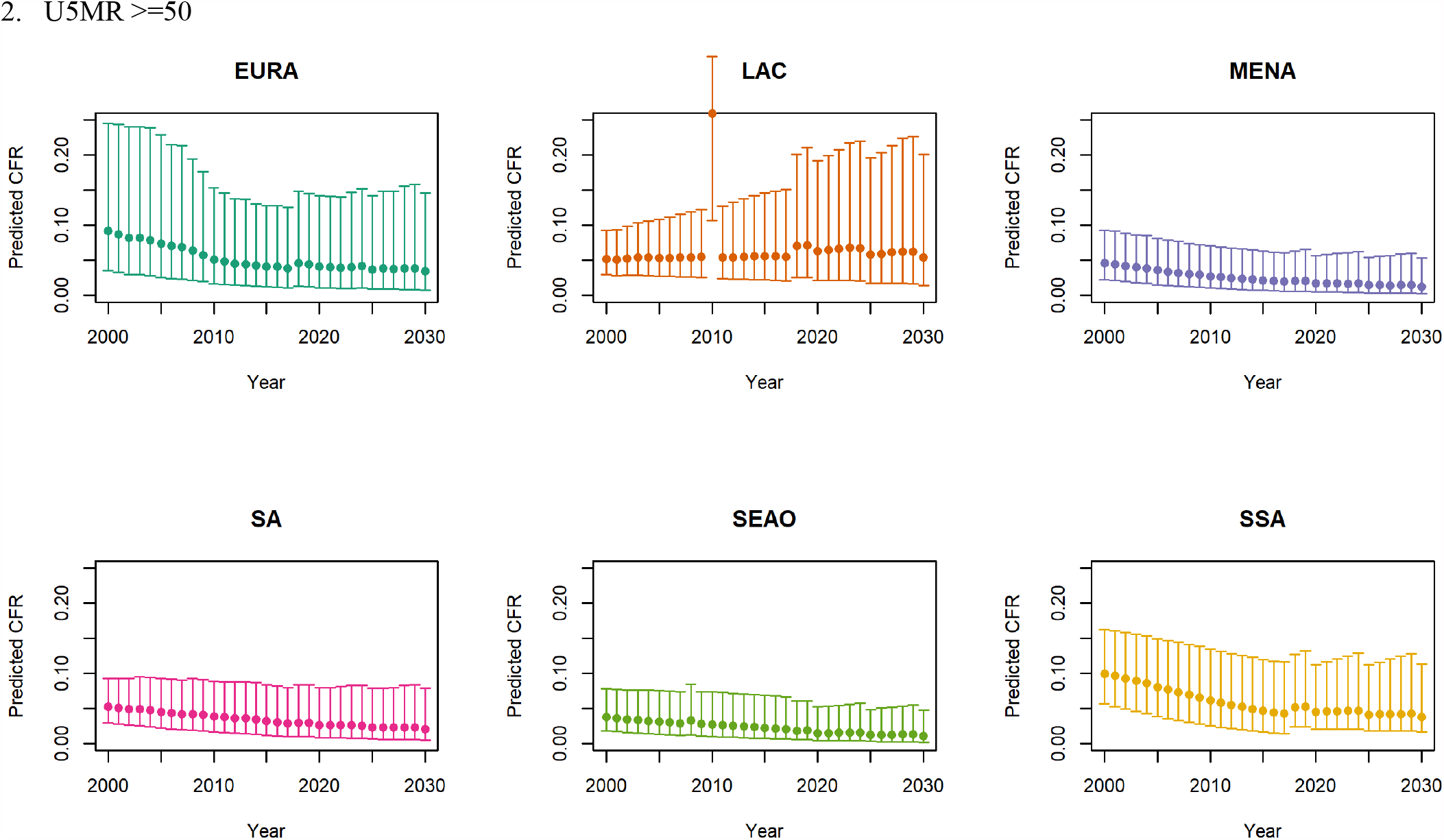

Note: The LAC region at >=50 U5MR only includes the country of Haiti, which experienced a significant shock to covariates due to the 2010 earthquake.

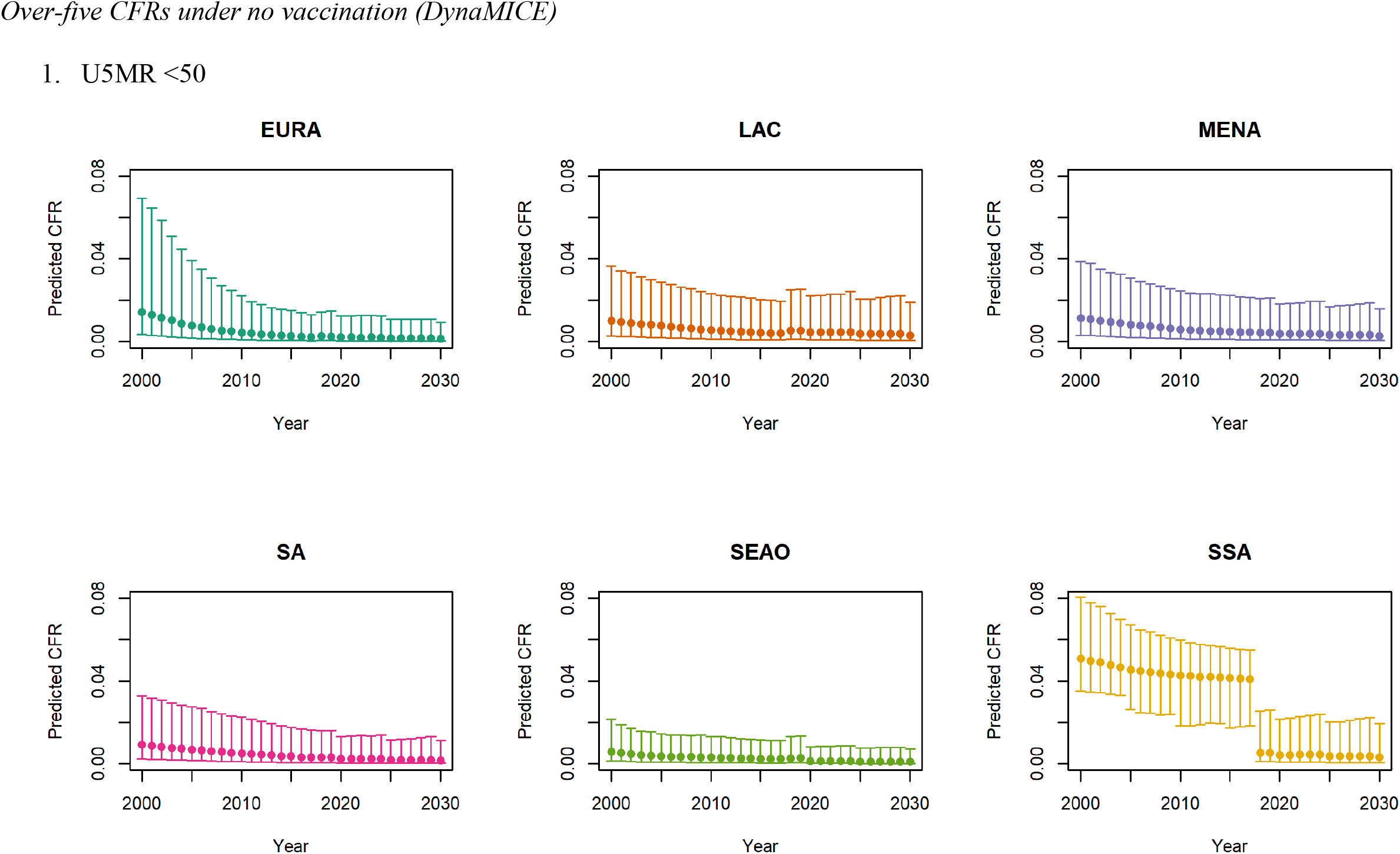
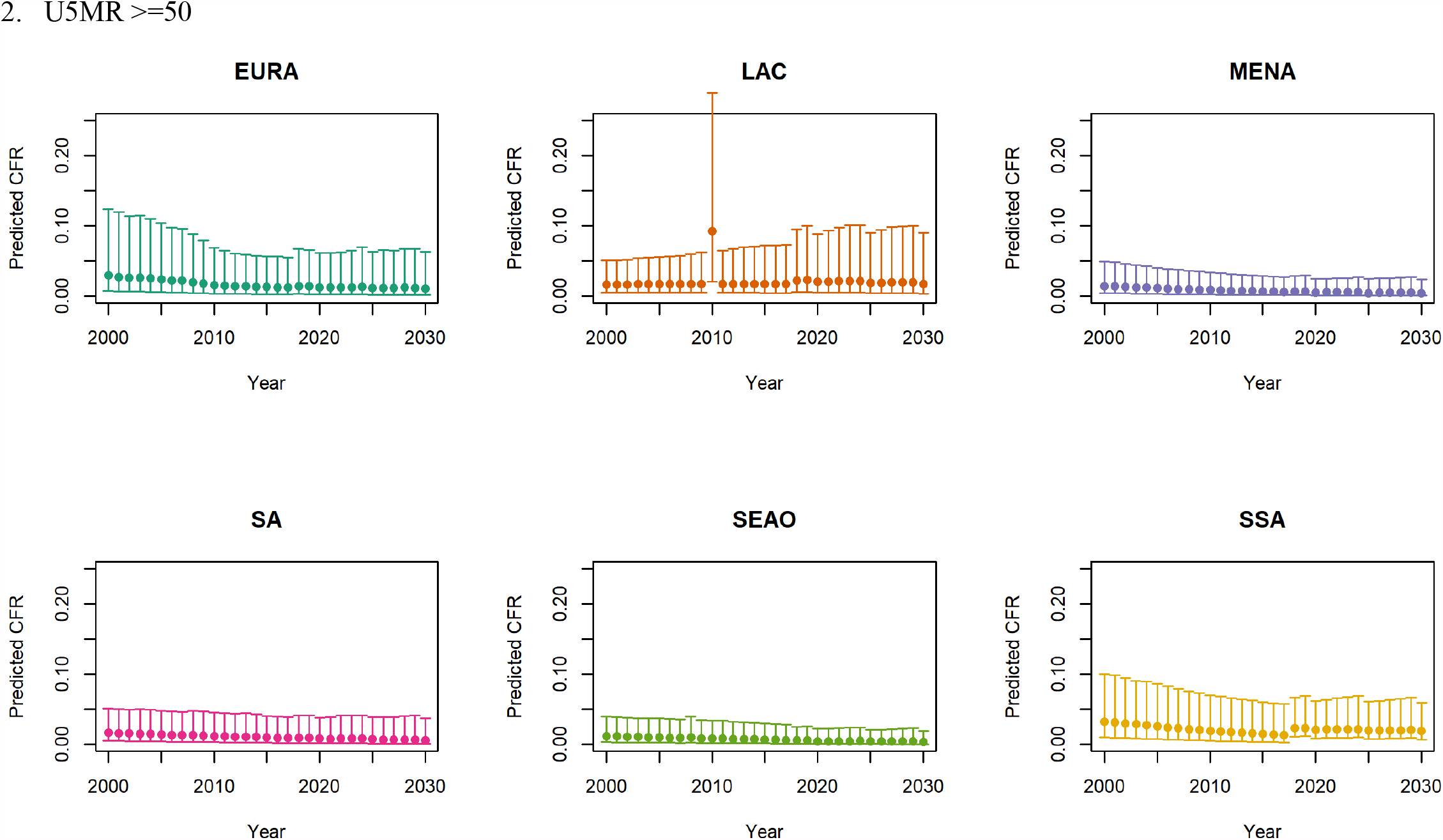

Note: The LAC region at >=50 U5MR only includes the country of Haiti, which experienced a significant shock to covariates due to the 2010 earthquake.

Note: EURA: Central Europe, Eastern Europe & Central Asia; LAC: Latin America & Caribbean; MENA: North Africa & Middle East; SA: South Asia; SEAO: South-East Asia, East Asia & Oceania; SSA: Sub-Saharan Africa; <50: less than 50 deaths per 1000 live births; >=50: greater than or equal to 50 deaths per 1000 live births.

## Footnotes

1 From Li, *et al*. (2021), supplementary appendix 2, with slight adaptations.

2 Verguet S, Johri M, Morris SK, Gauvreau CL, Jha P, Jit M. Controlling measles using supplemental immunization activities: a mathematical model to inform optimal policy. Vaccine 2015; 33:1291-6.

3 Verguet S, Jones EO, Johri M, et al. Characterizing measles transmission in India: a dynamic modeling study using verbal autopsy data. BMC Med 2017; 15.

4 Johri M, Verguet S, Morris SK, et al. Adding interventions to mass measles vaccinations in India. B World Health Organ 2016; 94:718-27.

5 Mossong J, Hens N, Jit M, et al. Social contacts and mixing patterns relevant to the spread of infectious diseases. Plos Med 2008; 5:381-91.

6 Uzicanin A, Zimmerman L. Field Effectiveness of Live Attenuated Measles-Containing Vaccines: A Review of Published Literature. J Infect Dis 2011; 204:S133-S48.

7 From Li, *et al*. (2021), supplementary appendix 2, with slight adaptations.

8 Finkenstadt BF, Grenfell BT. Time series modelling of childhood diseases. Applied Statistics 2000; 49:187-205.

9 Anderson RM, May RM. Infectious Disease of Humans: Dynamics and Control. Oxford Univerity Press, 1991.

10 Eilertson KE, Fricks J, Ferrari MJ. Estimation and prediction for a mechanistic model of measles transmission using particle filtering and maximum likelihood estimation. Statistics in Medicine 2019; 38:4146-58.

11 Simons E, Ferrari M, Fricks J, et al. Assessment of the 2010 global measles mortality reduction goal: results from a model of surveillance data. Lancet 2012; 379:2173-8.

12 Wolfson LJ, Grais RF, Luquero FJ, Birmingham ME, Strebel PM. Estimates of measles case fatality ratios: a comprehensive review of community-based studies. International Journal of Epidemiology 2009; 38:192-205.

